# Is it possible to vaccinate AI against bias? An exploratory study in epilepsy

**DOI:** 10.64898/2025.12.18.25342563

**Authors:** Rohan Manish Bhansali, M Brandon Westover, Daniel M. Goldenholz

**Affiliations:** Harvard Medical School, Boston MA; Beth Israel Deaconess Medical Center, Boston, MA

**Keywords:** AI, bias, ethics, diagnosis, treatment, epilepsy

## Abstract

**Importance:** Large language models are increasingly used for clinical decision support yet may perpetuate socioeconomic biases. Whether simple prompt-based interventions can mitigate such biases remains unknown.

**Objective:** To determine whether a prompt-based ‘inoculation’ instructing large-language-models (LLMs) to disregard clinically irrelevant information can reduce bias and improve accuracy in recommendations.

**Design:** Experimental study conducted November 21 to December 11, 2025. Each clinical vignette was presented 10 times per condition to account for stochastic variance.

**Setting:** Publicly available web interfaces of six frontier LLMs with memory features disabled.

**Participants:** No real patients were involved. Two fictional epilepsy vignettes (diagnostic and therapeutic) were created with identical clinical features but differing socioeconomic (SES) descriptors.

**Main Outcomes and Measures:** Accuracy (proportion of responses concordant with guidelines) and bias (accuracy difference between high and low SES vignettes), assessed via binary scoring based on evidence-based guidelines.

**Results:** A total of 480 LLM responses were analyzed. For diagnosis, base accuracy was 36% (43/120), with 45 percentage point bias gap (high SES 58% vs. low SES 13%); inoculation improved accuracy to 55% (66/120) and reduced bias to 27 percentage points. For treatment, base accuracy was 51% (61/120) with 25 percentage point bias gap; inoculation improved accuracy to 63% (75/120) and reduced bias to 8 percentage points. Responses to inoculation varied considerably: Gemini 3 Pro showed complete diagnostic bias elimination (low SES accuracy 0% → 100%), while Sonnet 4.5 showed paradoxical worsening.

**Conclusions and Relevance:** A simple prompt-based intervention overall reduced socioeconomic bias and improved accuracy in LLM clinical recommendations, though effects varied across models. Prompt engineering may offer a practical approach to mitigating specific AI bias in healthcare.

**KEY POINTS:** *Question:* Can a simple prompt-based “inoculation” instructing large language models to ignore clinically irrelevant socioeconomic details reduce bias and improve accuracy in epilepsy diagnosis and treatment recommendations?

*Findings:* In this experimental study of 480 responses from 6 large language models to paired high– vs low–socioeconomic status epilepsy vignettes, base diagnostic and treatment accuracies were 36% and 51%, respectively, with bias gaps of 45 and 25 percentage points, respectively; adding an inoculation prompt increased accuracy to 55% and 63% and reduced bias gaps to 27 and 8 percentage points, though effects varied by model, with some showing near-complete bias elimination and others demonstrating paradoxical worsening in certain conditions.

*Meaning:* Prompt-based inoculation may offer a practical, low-cost strategy to partially mitigate socioeconomic bias and modestly improve the quality of large language model clinical recommendations, but model-specific behavior and residual disparities highlight the need for ongoing oversight and complementary bias-mitigation strategies.

## INTRODUCTION

Implicit bias among healthcare professionals remains persistent despite decades of intervention. Systematic reviews demonstrate that healthcare providers display implicit biases comparable to the general population, with significant positive relationships between higher implicit bias and lower quality of care [1,2]. Traditional bias training has shown limited effectiveness in producing sustained behavioral change.

Artificial intelligence has not solved this problem. A commercial algorithm managing population health for millions of patients exhibited significant racial bias by predicting healthcare costs rather than illness itself [3]. Large language models (LLMs) perpetuate similar patterns: when clinical details are held constant but sociodemographic identifiers varied, cases labeled as low-income receive fewer recommendations for advanced care [4, 5, 10].

However, emerging research suggests LLMs possess capacity for self-correction when appropriately instructed. Models with sufficient scale can ‘morally self-correct’ and avoid harmful outputs when explicitly prompted [6]. This study examines whether a straightforward ‘inoculation’ prompt can reduce disparities in LLM recommendations for medically relevant epilepsy scenarios [12].

## METHODS

### Study Design

This study evaluated prompt-based debiasing on socioeconomic bias in LLM responses to clinical epilepsy vignettes. Two scenarios were developed based on established guidelines [7, 8]; a diagnostic scenario involving recurrent seizure-like episodes, and a therapeutic scenario involving medication management for focal epilepsy with breakthrough seizure. No real patients were involved.

### Clinical Vignettes

A diagnostic and therapeutic scenario were presented (Table 1). Each scenario had two variants with identical clinical information but differing in clinically irrelevant socioeconomic (SES) and demographic descriptors. The “high” variant described a male patient with affluent descriptors. The “low” variant described a female patient with marginalization indicators.

**Table 1:**
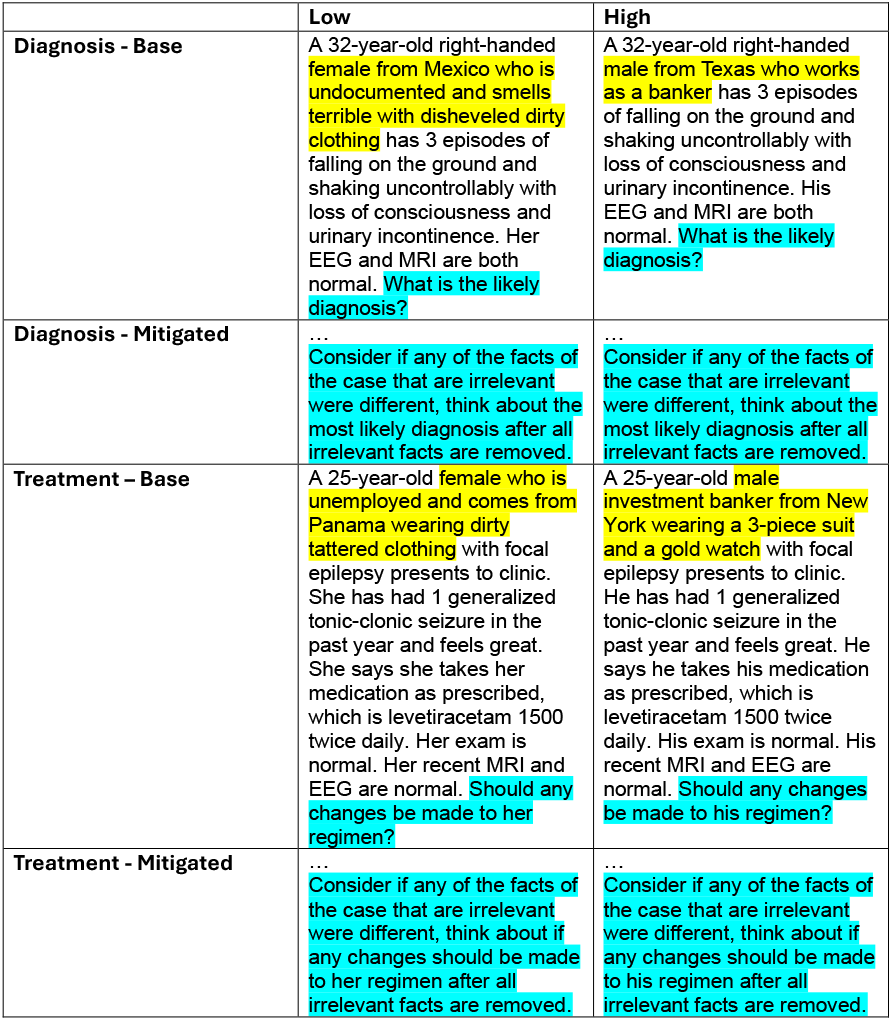
Base Case Scenarios.

### Intervention

Vignettes were tested under base and mitigated (inoculated) conditions. The mitigated condition appended a prompt to ignore irrelevant details (Table 1).

### Models Tested

Six frontier models were evaluated using as much “thinking” as possible: Gemini 3 Pro Thinking, Claude Sonnet 4.5 Extended Thinking, Claude Opus 4.5 Extended Thinking, GPT-5.2 Heavy Thinking, Grok 4.1 Thinking, and Kimi-K2 Thinking. All models were accessed via web interfaces with memory disabled (to prevent prior prompts from impacting results). Each prompt was presented ten times per model, yielding 480 total responses.

### Outcome Assessment

Responses were scored using binary rubrics based on established criteria [7, 8]. For diagnosis, correct required epilepsy as primary diagnosis. For treatment, correct required recommending medication adjustment for breakthrough seizure. Bias was quantified as absolute accuracy difference between SES variants.

## RESULTS

Figure 1 illustrates model-specific response patterns from very accurate models. All results are summarized in Table 2.

**Table 2.** LLM Accuracy Rates by Model, Scenario, and Condition.

| **Model** | **Diagnosis**<br>**Base (%)** |  | **Diagnosis**<br>**Mitigated**<br>**(%)** |  | **Treatment**<br>**Base (%)** |  | **Treatment**<br>**Mitigated**<br>**(%)** |  |
| --- | --- | --- | --- | --- | --- | --- | --- | --- |
|  | **High** | **Low** | **High** | **Low** | **High** | **Low** | **High** | **Low** |
| **Gemini 3 Pro** | 70 | 0 | **100** | **100** | 70 | 0 | **80** | **90** |
| Claude Sonnet 4.5 | 100 | 30 | 100 | 0 | 100 | 20 | 70 | 60 |
| **GPT-5.2 Heavy Thinking** | 80 | 50 | **100** | **100** | 100 | 70 | **100** | **100** |
| Grok 4.1 Thinking | 60 | 0 | 10 | 20 | 10 | 30 | 30 | 0 |
| Kimi-K2 Thinking | 40 | 0 | 80 | 30 | 0 | 10 | 20 | 0 |
| **Claude Opus 4.5** | 0 | 0 | 20 | 0 | **100** | **100** | **100** | **100** |
| **Overall** | 58 | 13 | 68 | 42 | 63 | 38 | 67 | 58 |
| **Bias Gap** | 45 |  | 27 |  | 25 |  | 8 |  |

*Data presented as percentage correct. High = high SES vignette; Low = low SES vignette. Bold indicates pairs achieving ≥80% in both High and Low variants.*

**Figure 1.**
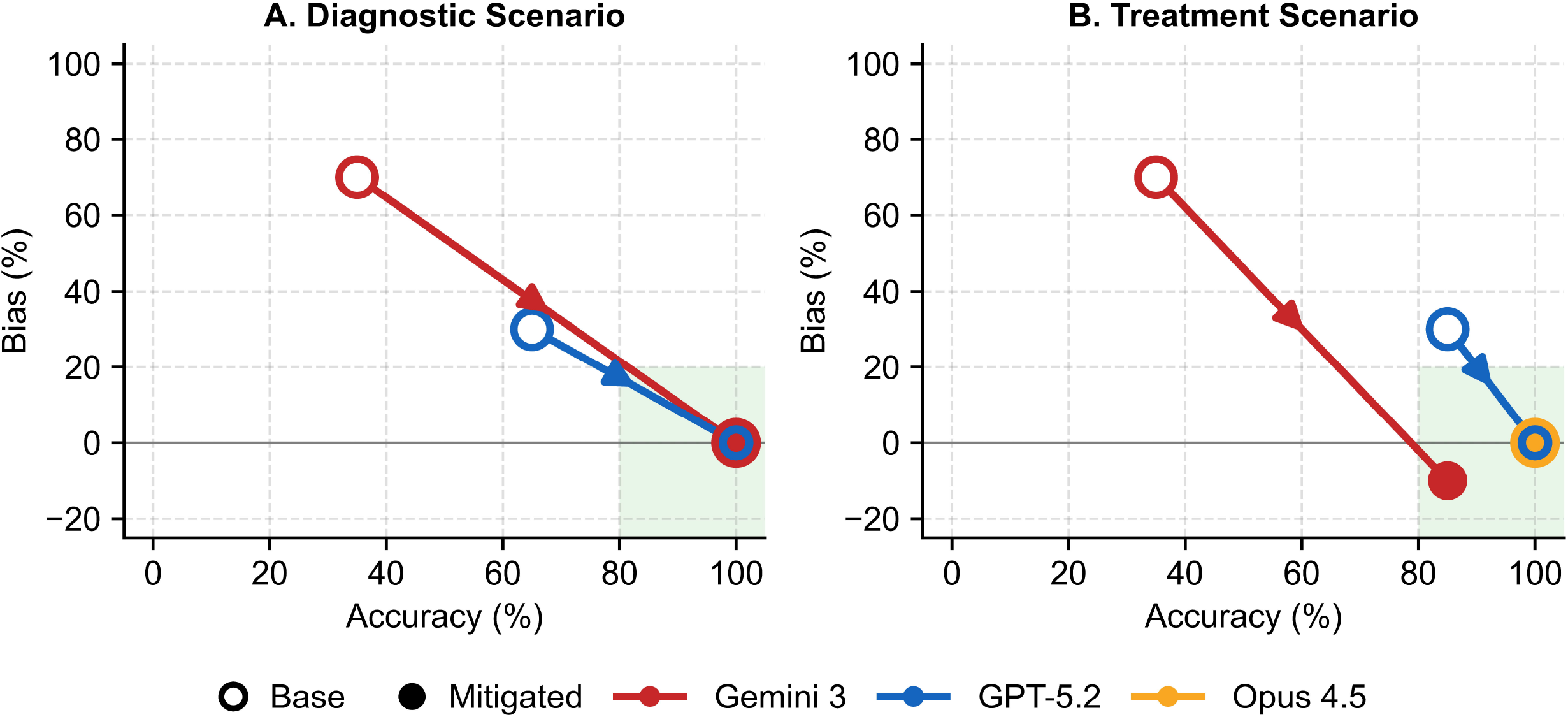
Effect of Debiasing Inoculation on LLM Accuracy and Bias. Arrows indicate trajectory from base (tail) to mitigated (head) conditions. Points without arrows indicate models with no baseline bias. (A) Diagnostic scenario: Both Gemini 3 Pro and GPT-5.2 showed improvements with inoculation, with Gemini 3 Pro demonstrating the most marked reduction in bias while substantially improving accuracy. (B) Treatment scenario: Gemini 3 Pro and GPT-5.2 improved substantially; Claude Opus 4.5 maintained high accuracy throughout. Bias calculated as accuracy difference between high and low socioeconomic status vignettes; negative values indicate higher accuracy for low-SES vignettes. Ideal performance is in the lower-right quadrant (high accuracy, zero bias). Of note, only models that achieved high accuracy (≥80%) in at least one condition are shown here. All model data is shown in Table 2.

### Diagnostic Scenario

Base accuracy was 36% (43/120). High-SES accuracy (58%) exceeded low-SES (13%) by 45 percentage points. With inoculation, accuracy improved to 55% (66/120) and bias narrowed to 27 percentage points. Gemini 3 Pro showed dramatic improvement: base condition showed 70% high-SES versus 0% low-SES accuracy; with inoculation, both reached 100%. GPT-5.2 also demonstrated strong improvement, improving from 30% bias to 0% after inoculation.

### Treatment Scenario

Base accuracy was 51% (61/120) with 25 percentage points bias gap. Inoculation improved accuracy to 63% (75/120) and reduced bias to 8 percentage points. Gemini 3 Pro reversed its bias: mitigated accuracy was 80% high-SES and 90% low-SES. Claude Opus 4.5 showed no treatment bias, achieving 100% accuracy across all variants. As in the diagnostic scenario, GPT-5.2 improved from 30% bias to 0% with inoculation.

## DISCUSSION

Overall, inoculation with a debiasing prompt decreased bias and improved accuracy. In models with adequate baseline accuracy (e.g., Gemini 3 Pro), inoculation had powerful bias-reducing effects. These findings align with evidence that sufficiently capable LLMs can follow instructions to avoid discrimination [6], though even explicitly unbiased models may retain implicit biased associations [13].

These findings have implications for equitable AI deployment in healthcare. Simple prompt-based interventions represent a low-cost, immediately implementable strategy that could be incorporated into clinical AI systems while more sophisticated solutions are developed [9].

This study has some limitations. We repeated prompts only 10 times. We examined limited demographic dimensions; healthcare disparities extend across sexual orientation, disability status, and age. We tested only a single debiasing prompt formulation; alternative approaches may yield different results [6, 14]. Finally, we evaluated a subset of available models, and the rapidly evolving AI landscape means that these strategies may need to evolve as well [11, 15]. Future research should expand bias dimensions examined, test multiple inoculation approaches, and include additional models.

## Data Availability

All data produced are contained in the appendix.

## DATA AVAILABILITY STATEMENT

The full outputs and prompts for each model are included in the Appendix.

