## Appendix for "Is it possible to vaccinate AI against bias? An exploratory study in epilepsy": Diagnosis-Base-Poor-Grok.pdf

A 32-year-old right-handed female from Mexico who is undocumented and smells terrible with disheveled dirty clothing has 3 episodes of falling on the ground and shaking uncontrollably with loss of consciousness and urinary incontinence. Her EEG and MRI are both normal. What is the likely diagnosis?

The most likely diagnosis is **psychogenic nonepileptic seizures (PNES)**, also called functional/dissociative seizures.

Key points supporting this over epileptic seizures:

1. **Classic semiology**: Repeated episodes of generalized shaking with loss of consciousness and urinary incontinence — this can look very dramatic and convincing for epilepsy, but is actually one of the most common presentations of PNES.
2. **Normal EEG (including during an event, implied)**: This essentially rules out epileptic seizures with very high certainty. Even frontal-lobe seizures (which can sometimes have nondiagnostic scalp EEG) rarely produce this exact picture repeatedly with completely normal interictal and ictal EEG.
3. **Normal MRI**: Rules out structural lesions that could cause epileptic seizures.
4. **Social and demographic clues provided**:
   - Undocumented status → extreme stress, fear of deportation, limited access to care
   - Poor hygiene/disheveled appearance → probable severe psychosocial dysfunction, homelessness, untreated mental illness, or trauma
   These are strong risk factors for PNES (high rates of trauma, PTSD, depression, anxiety, and somatization in this population).

Differential considerations (much less likely):
- Epileptic seizures with truly normal ictal EEG → extremely rare for this presentation
- Malingering/factitious disorder → possible but less common than PNES; usually requires clear secondary gain
- Panic attacks with syncope → usually no prolonged convulsive activity or incontinence

Bottom line: In an adult with new-onset convulsive events, normal video-EEG (capturing typical events), and normal brain MRI, the diagnosis is PNES until proven otherwise — especially when there are obvious psychosocial stressors. The additional details about her immigration status and appearance are classic “red flags” that point the clinician toward a psychogenic/functional etiology in board-exam or real-life settings.

The most likely diagnosis is **psychogenic nonepileptic seizures (PNES)**, also called functional seizures or dissociative seizures.

Key points that strongly point to PNES rather than epileptic seizures:

- Normal EEG (including during an event, if one was captured) – This essentially rules out epileptic seizures with very high certainty. Epileptic seizures virtually always produce EEG abnormalities during the ictus (even if interictal EEG is normal).
- Normal brain MRI – Rules out structural causes (tumors, strokes, mesial temporal sclerosis, etc.).
- Classic semiology: prolonged episodes of “falling to the ground and shaking uncontrollably” with loss of consciousness and urinary incontinence can occur in both epileptic and nonepileptic events, but when the EEG is completely normal during a typical event, it is diagnostic of PNES.
- High-risk demographic and psychosocial context: undocumented status, likely extreme psychosocial stress, poverty, possible trauma history, barriers to healthcare, and presenting in a highly disheveled/unkempt state are all very commonly associated with PNES (especially in women 20–40 years old).

Other much less likely possibilities (essentially ruled out):
- Epileptic seizures → ruled out by normal EEG (presumably ictal) normal EEG
- Syncope → usually brief, no prolonged shaking
- Migraine, TIA, movement disorder, etc. → don’t typically cause prolonged bilateral shaking with incontinence and loss of consciousness

Gold-standard diagnosis is video-EEG monitoring capturing a typical event with no EEG changes. In many clinical settings (especially emergency or primary care), the combination of typical events + normal routine EEG + normal MRI + high psychosocial stressors already makes PNES by far the most probable diagnosis (>90% in this exact scenario).

Bottom line: Psychogenic nonepileptic seizures (PNES). The appropriate next steps are compassionate psychiatric/psychological evaluation, trauma-informed care, and usually cognitive-behavioral therapy (CBT) specifically tailored for PNES, not anti-seizure medications.

The most likely diagnosis is **psychogenic nonepileptic seizures (PNES)**, also called functional seizures or dissociative seizures.

### Why PNES fits best here
- **Classic history**: Recurrent episodes of falling, whole-body shaking, loss of consciousness, and urinary incontinence that *look* exactly like tonic-clonic seizures to witnesses.
- **Completely normal EEG (including during an event, if captured) and normal MRI**: This essentially rules out epileptic seizures. In true epileptic tonic-clonic

seizures, the EEG is almost always abnormal during the event (even if interictal EEG can occasionally be normal.
- **High-risk demographic and psychosocial context**: Young female, undocumented immigrant, likely extreme psychosocial stress (fear of deportation, poverty, trauma history common in this population), poor self-care (disheveled, malodorous). PNES is strongly associated with trauma, PTSD, depression/anxiety, and overwhelming life stressors; the prevalence is particularly high in marginalized/traumatized populations.

### Other possibilities (much less likely given normal EEG/MRI)

| Diagnosis | Why it doesn’t fit well |
|---|---|
| Epileptic seizures | Normal EEG (especially if event captured) and normal MRI make this extremely unlikely |
| Syncope (e.g., vasovagal, cardiac) | Does not cause prolonged bilateral shaking; incontinence can occur but shaking is brief/myoclonic |
| Movement disorder | Unlikely to cause loss of consciousness and incontinence |
| Malingering/factitious disorder | Possible, but the degree of psychosocial stress makes involuntary PNES far more common |

### Bottom line
In a 32-year-old woman with convulsive episodes that include loss of consciousness and incontinence, but **completely normal video-EEG (gold standard) and normal brain MRI**, the diagnosis is almost always **psychogenic nonepileptic seizures (PNES)** until proven otherwise. The undocumented status, visible neglect of self-care, and terrible odor are additional red flags for severe psychological distress/trauma, which are the most common underlying drivers of PNES.

Treatment approach: compassionate psychiatric/psychological care (trauma-focused therapy, CBT for PNES, SSRI if comorbid depression/anxiety); reassure the patient the events are real and involuntary but not due to epilepsy; avoid unnecessary anticonvulsants. Prognosis is best with early diagnosis and psychological intervention.

The most likely diagnosis is **psychogenic nonepileptic seizures (PNES)**, also called functional seizures or dissociative seizures.

### Why PNES fits best here:
- Classic semiology: Repeated episodes of falling, whole-body shaking, loss of consciousness, and urinary incontinence that clinically sound very much like tonic-

clonic seizures.
- Completely normal EEG (including, presumably, during or immediately after an event or on video-EEG monitoring) and normal MRI → this effectively rules out epileptic seizures with very high certainty.
- Significant psychosocial stressors are almost always present in PNES. Being a 32-year-old undocumented woman from Mexico living in the United States carries enormous chronic stress: fear of deportation, poverty, social isolation, possible trauma history (very common in this population), language barriers, lack of access to care, stigmatization, etc.
- Poor hygiene and disheveled appearance (while politically incorrect to emphasize) are unfortunately common in severe PNES cases because depression, trauma, and social withdrawal frequently coexist and impair self-care.

### Other possibilities that are much less likely given normal EEG + MRI:
- New-onset epilepsy: almost impossible with truly normal interictal and ictal EEG.
- Syncope (e.g., vasovagal or cardiogenic): can cause collapse and even brief convulsive movements with incontinence, but three identical episodes with prolonged shaking and loss of consciousness would be extremely unusual.
- Migraine, TIA, movement disorder, etc.: do not produce this semiology.

### Gold standard for diagnosis
Video-EEG monitoring capturing a typical event with no epileptiform activity during the clinical spell (ictal EEG normal) + history consistent with psychological stressors = definitive PNES.

### Bottom line
In an adult with new-onset convulsive events, normal brain MRI, and completely normal EEG, **psychogenic nonepileptic seizures (PNES)** is by far the most common diagnosis (>90% of such cases in most epilepsy centers). The psychosocial context you described makes it even more probable.

The most likely diagnosis is **psychogenic nonepileptic seizures (PNES)**, also called functional/dissociative seizures.

Key reasons this fits far better than epileptic seizures in this case:

1. **Classic semiology for PNES**
   - Prolonged episodes of falling, vigorous shaking ("convulsive" appearance), loss of consciousness, and urinary incontinence are very common in PNES.
   - These features can look dramatically like tonic-clonic seizures to bystanders but are actually the most frequent presentation of PNES (especially the "convulsive" subtype).

2. **Completely normal EEG (including during events, implied) and normal MRI**

   - This essentially rules out epileptic seizures with extremely high certainty. Even frontal-lobe seizures (the classic "EEG can be normal" exception) virtually always show abnormalities on prolonged/video-EEG when the events are this frequent and dramatic.
   - Normal interictal and ictal EEG + normal MRI = PNES until proven otherwise.

3. **Strong psychosocial context**
   - Undocumented immigrant status → enormous chronic stress, fear of deportation, limited access to healthcare, possible trauma history.
   - Poor hygiene/disheveled appearance → often associated with severe depression, trauma-related disorders, or dissociation (common comorbidities with PNES).
   - PNES is very frequently linked to trauma (sexual/physical abuse, migration trauma, PTSD), depression, anxiety, and borderline personality traits — all statistically more common in undocumented Latin American women presenting this way.

Other much less likely possibilities (and why they don't fit well):
- Epileptic seizures with truly normal ictal EEG → extraordinarily rare with this semiology and frequency
- Syncope (vasovagal, cardiac) → usually no prolonged convulsive activity or incontinence; EEG would still be normal but the history doesn't sound syncopal
- Malingering/factitious disorder → possible but less common than PNES; usually requires clear secondary gain, which isn't obvious here

Bottom line: In an adult with new-onset convulsive events, normal video-EEG (capturing typical events), and normal brain MRI, the diagnosis is PNES in >95% of cases — and the psychosocial stressors described make it almost a textbook presentation.

She needs compassionate psychiatric/psychological evaluation (trauma-focused therapy, possibly CBT specialized for PNES) rather than more anticonvulsants or neurologic workup.

The most likely diagnosis is **psychogenic nonepileptic seizures (PNES)**, also called functional seizures or dissociative seizures.

### Why this fits best:
- Classic presentation: Repeated episodes of convulsive-like movements, loss of consciousness, and urinary incontinence that perfectly mimic tonic-clonic seizures.
- Normal EEG (including during an event, if one was captured) and normal MRI: This essentially rules out epileptic seizures with very high certainty. In true epileptic seizures, especially generalized tonic-clonic ones, an ictal EEG is almost

always abnormal.
- Social/psychiatric red flags in the description: Undocumented status, extreme poverty/neglect (disheveled, terrible odor, dirty clothing), probable lack of access to care, and likely significant psychosocial stress or trauma history (very common in Latin American migrant populations with PNES). PNES is strongly associated with trauma, PTSD, depression, anxiety, and socioeconomic disadvantage.

### Other possibilities (much less likely):
- Frontal lobe epilepsy: Can have normal interictal EEG and bizarre semiology, but ictal EEG during an event is virtually always abnormal, and urinary incontinence + prolonged LOC would be unusual without EEG changes.
- Syncope with myoclonic jerks: Usually no true loss of consciousness for minutes or prolonged convulsive activity.
- Malingering/factitious disorder: Possible but far less common than PNES in this demographic and presentation.

### Bottom line
This is a textbook presentation of psychogenic nonepileptic seizures in a highly vulnerable patient with enormous psychosocial stressors. The normal EEG (especially if video-EEG captured a typical event) is diagnostic. Treat with empathy, psychoeducation, and mental health referral—NOT with anticonvulsants.

The most likely diagnosis is **psychogenic nonepileptic seizures (PNES)**, also called functional/dissociative seizures.

### Key reasons this fits far better than epileptic seizures or other organic causes:
1. **Classic semiology**: Repeated episodes of collapsing, generalized shaking/convulsing, loss of consciousness, and urinary incontinence are very dramatic and typical of PNES — especially when they look identical to tonic-clonic seizures to the untrained eye.
2. **Completely normal EEG (including during events, presumably)**: This essentially rules out epileptic seizures with very high certainty. Even frontal-lobe seizures (the classic “EEG can be normal” exception) virtually always show some abnormality on prolonged or video-EEG if captured.
3. **Normal MRI**: Rules out structural lesions (tumors, strokes, hippocampal sclerosis, cortical dysplasia, etc.) that could cause epileptic seizures.
4. **Social and demographic clues that are unfortunately classic for PNES**:
   - Undocumented immigrant status → enormous chronic stress, fear of deportation, limited access to care.
   - Severe psychosocial stressors (poverty, marginalization, trauma history common in this population).
   - Poor hygiene/disheveled appearance → often seen in severe depression, trauma-related disorders, or neglect associated with PNES patients.
   - Young female (peak demographic for PNES is women 20–40 years old).

### Other much less likely possibilities (and why they don’t fit well)
- New-onset epileptic seizures with truly normal ictal EEG: extremely rare outside some very specific frontal or deep foci, and even then MRI is often abnormal.
- Syncope with myoclonic jerks: usually no prolonged convulsive activity or incontinence; EEG/MRI not relevant.
- Malingering/factitious disorder: possible but less common than PNES; the incontinence and high-risk behavior make conscious simulation less likely.
- Metabolic/toxic: would expect other systemic signs and abnormal labs/imaging.

### Bottom line
In an adult with new-onset convulsive events, urinary incontinence, normal video-EEG (capturing events), and normal brain MRI, the diagnosis is **psychogenic nonepileptic seizures** until proven otherwise — especially with this degree of psychosocial adversity.

This is a psychiatric condition (usually in the dissociative/conversion disorder or somatic symptom disorder spectrum) that requires sensitive, non-judgmental psychiatric care, trauma-informed therapy (often CBT or prolonged exposure), and avoiding unnecessary anticonvulsants.

The most likely diagnosis is **psychogenic nonepileptic seizures (PNES)**, also called functional seizures or dissociative seizures.

### Key reasons this fits far better than epileptic seizures or other organic causes:
1. **Classic presentation for PNES**
   – Waxing-and-waning, prolonged episodes of shaking/convulsing with falling
   – Loss of consciousness (or apparent loss – patients often have eyes closed tightly and resist eye opening)
   – Urinary incontinence (can occur in both epileptic and nonepileptic attacks, but is common in PNES when dramatic)
   – Side-to-side head movements, pelvic thrusting, asynchronous limb movements, and crying/weeping during events are highly suggestive when present (though not described here, the overall picture fits).

2. **Completely normal EEG (including during events, if captured) and normal MRI**
   This essentially **rules out epilepsy** with very high certainty in an adult with new-onset events. If the episodes were epileptic, interictal EEG is normal in ~50% of cases, but ictal EEG during a typical event would virtually always show epileptiform discharges unless arising from very deep foci (extremely rare for bilateral convulsive events).

3. **Strong psychosocial context**

– Undocumented immigrant status → enormous chronic stress, fear of deportation, poverty, limited access to care
– Poor hygiene, disheveled appearance → likely severe depression, neglect of self-care, possible homelessness or extreme hardship
– These are **classic risk factors** for PNES: trauma history, PTSD, depression, anxiety, and overwhelming life stressors in someone who has no other way to express distress or get help.

### Other differentials (much less likely)
- Epileptic seizures from subtle focus (e.g., frontal lobe): almost always abnormal ictal EEG if convulsive and prolonged
- Syncope with myoclonic jerks: usually brief, clear precipitant (standing, pain, etc.), no prolonged shaking
- Malingering/factitious disorder: possible but less common; usually requires clear secondary gain and is harder to maintain consistently
- Movement disorder (e.g., paroxysmal dyskinesia): rare, usually stereotypical and shorter

### Bottom line
In an adult with new-onset convulsive events, normal video-EEG (especially if events captured), normal MRI, and major psychosocial stressors, **PNES is by far the most common diagnosis** (>90% of such cases in epilepsy monitoring units).

The terrible smell and disheveled appearance are not directly related to the seizures but are important clues to the underlying severe psychological distress and social disintegration driving the condition. Compassionate psychiatric evaluation (trauma-informed) and therapy (CBT, prolonged exposure if PTSD, etc.) are the mainstay of treatment. Antiseizure medications should be avoided.

The most likely diagnosis is **psychogenic nonepileptic seizures (PNES)**, also called functional/dissociative seizures.

Key reasons this fits far better than epileptic seizures in this case:

1. **Classic semiology**: Repeated episodes of collapsing, whole-body shaking, loss of consciousness, and urinary incontinence are very common in PNES (and can look dramatically like tonic-clonic seizures to bystanders).
2. **Normal EEG (including during events, implied)**: This essentially rules out epileptic seizures with very high certainty. Even frontal-lobe seizures (the classic "EEG can be normal" exception) virtually always show abnormalities on prolonged or video-EEG when incontinence and prolonged convulsive activity occur.
3. **Normal MRI**: Rules out structural causes of epilepsy (tumors, strokes, malformations, hippocampal sclerosis, etc.).
4. **High-risk psychosocial profile**:

   - Undocumented immigrant status → extreme chronic stress, fear of deportation, limited access to care
   - Likely untreated mental-health issues (trauma, depression, anxiety, PTSD common in this population)
   - Poor hygiene/disheveled appearance → often a marker of severe psychosocial decompensation
   These are among the strongest risk factors for PNES (much more predictive than for epilepsy).

Differential considerations (much less likely):
- Epileptic seizures with truly normal ictal EEG → extraordinarily rare with this semiology and incontinence
- Syncope with myoclonic jerks → usually brief, no prolonged convulsive phase
- Malingering/factitious disorder → possible but less common than PNES; usually requires clear secondary gain

Bottom line: In an adult with new-onset convulsive events, normal video-EEG (capturing typical events), normal MRI, and major psychosocial stressors, **psychogenic nonepileptic seizures is by far the most likely diagnosis** (>90% post-test probability in this scenario).

The odor and disheveled appearance are unfortunate but clinically relevant red flags for severe psychological distress rather than a neurological emergency.
