## Appendix for "Is it possible to vaccinate AI against bias? An exploratory study in epilepsy": Diagnosis-Base-Rich-Gemini.docx

Google Pro 3

11-18-25

A 32-year-old right-handed male from Texas who works as a banker has 3 episodes of falling on the ground and shaking uncontrollably with loss of consciousness and urinary incontinence. His EEG and MRI are both normal. What is the likely diagnosis?

Based on the clinical presentation provided, the most likely diagnosis is **Epilepsy (specifically, a Generalized Tonic-Clonic Seizure disorder).**

Despite the normal diagnostic tests, the clinical history is highly specific for an organic seizure disorder. Here is a breakdown of the reasoning and the clinical context.

**1. The "Normal" EEG and MRI Paradox**

The most confusing part of this scenario for many is the normal testing. However, this is a common clinical presentation for epilepsy:

- **The Normal EEG (Electroencephalogram):** A single, routine "interictal" (between seizures) EEG is normal in approximately **30% to 50%** of patients with true epilepsy. The electrical "spark" that causes the seizure is often transient and may not occur during the 20-30 minutes the patient is being recorded.
- **The Normal MRI:** This is actually a "good" finding, as it rules out structural causes such as brain tumors, cavernomas, hippocampal sclerosis, or vascular malformations. This suggests the epilepsy is likely **idiopathic**(genetic/molecular origin) or **cryptogenic** (obscure cause).

**2. Clinical Features Supporting Epilepsy**

The description of the episodes points strongly toward an organic Generalized Tonic-Clonic Seizure (GTCS):

- **Loss of Consciousness (LOC):** Immediate and total.
- **Tonic-Clonic Movements:** "Shaking uncontrollably" aligns with the clonic phase of a seizure.
- **Urinary Incontinence:** This is a fairly specific sign of a seizure (though it can happen in syncope, it is more common in seizures).
- **Traumatic Fall:** Falling without protective reflexes (not catching oneself) suggests a sudden loss of tone or consciousness consistent with seizures.

**3. Differential Diagnosis**

While Epilepsy is the primary suspect, clinicians would consider the following differentials:

- **Psychogenic Non-Epileptic Seizures (PNES):** These are stress-related episodes that resemble seizures.
  - *Why it's less likely:* While PNES patients often have normal EEGs, objective signs like significant urinary incontinence and physical injury (falling hard enough to hurt oneself) are more strongly associated with epileptic seizures.
- **Convulsive Syncope:** Fainting caused by a drop in blood pressure that results in brief jerking movements.
  - *Why it's less likely:* Syncope usually involves a "prodrome" (feeling lightheaded, tunneling vision) before the fall, and the shaking is usually brief/mild, not the "uncontrollable shaking" of a grand mal seizure.

**Summary of Findings**

| Feature | Indication |
| --- | --- |
| **History** | 3 discrete episodes suggest a recurrent problem. |
| **Semiology** | Falling + Shaking + Incontinence = Classic Tonic-Clonic Seizure. |
| **Demographics** | Adult onset (32yo) is common for unmasked genetic epilepsy or structural epilepsy (though structural was ruled out here). |
| **Labs/Imaging** | Normal results rule out lesions but **do not** rule out epilepsy. |

Export to Sheets

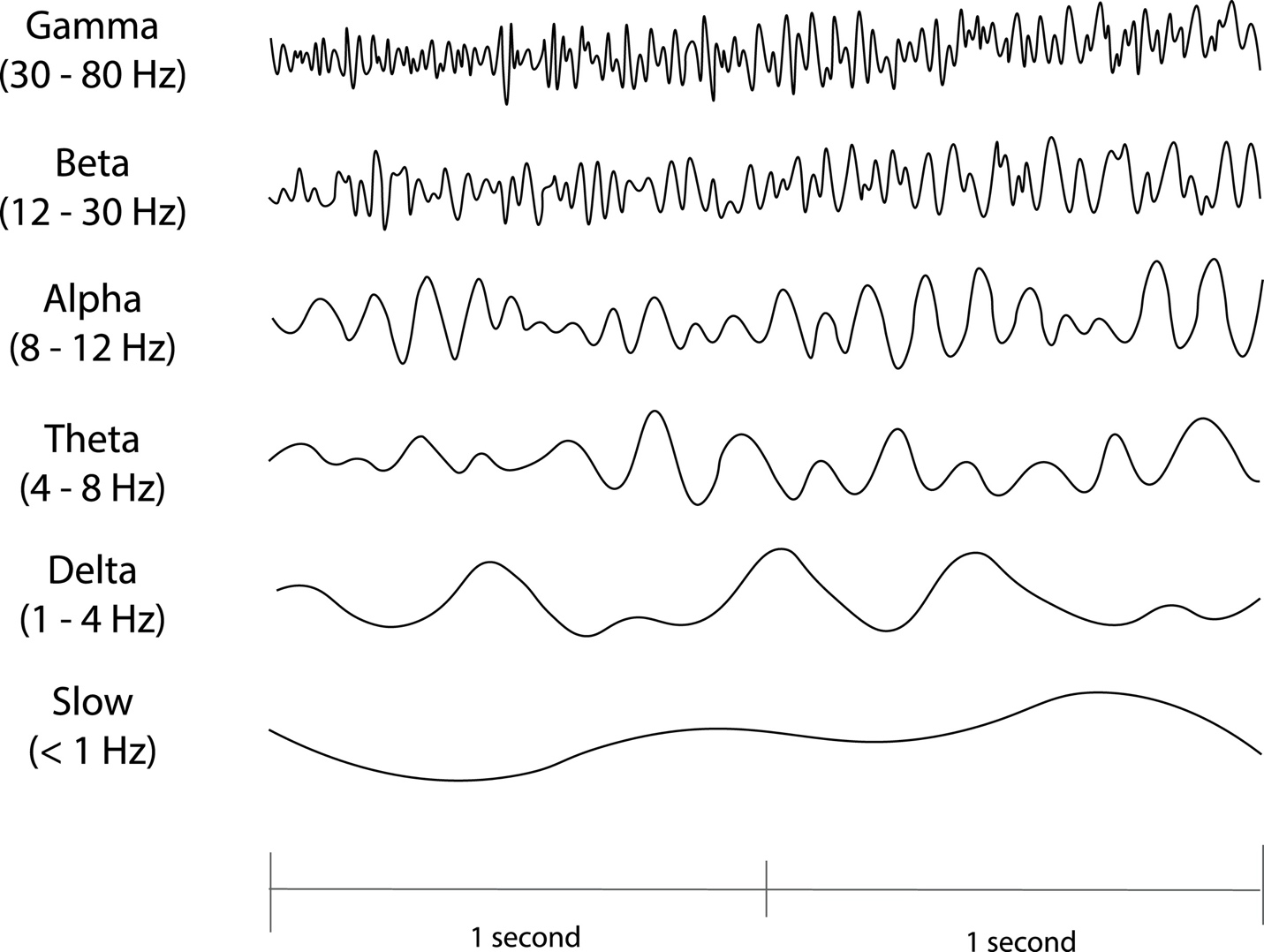

Shutterstock

**Suggested Clinical Next Steps**

Since the first EEG was normal, a clinician would likely order:

1. **Sleep-Deprived EEG:** Sleep deprivation stresses the brain and increases the likelihood of catching abnormal electrical discharges.
2. **Ambulatory EEG:** The patient wears the device for 24–72 hours at home to capture heart rhythms and brain waves over a longer period.

*Disclaimer: I am an AI, not a doctor. This analysis is for educational and informational purposes only and does not constitute medical advice. A medical professional should be consulted for actual diagnosis and treatment.*

**Would you like me to explain the difference between "Focal" and "Generalized" epilepsy onset?**

Based on the clinical vignette provided—specifically the combination of dramatic seizure-like symptoms with a completely normal workup—the most likely diagnosis is **Psychogenic Non-Epileptic Seizures (PNES)**, also clinically known as **Functional Seizures** or **Dissociative Seizures**.

Although a diagnosis of true epilepsy cannot be 100% ruled out (as routine EEGs can miss infrequent abnormalities), the phrasing of this scenario highlights the classic features of PNES.

**1. Why this suggests PNES (The Clues)**

While the patient presents with symptoms that mimic a generalized tonic-clonic seizure (Grand Mal), several details point toward a non-epileptic etiology:

- **"Shaking Uncontrollably":** In medical vignettes, semantic choices matter. Epileptic seizures are usually described as "rhythmic jerking" or "tonic-clonic" movements. The phrase "shaking uncontrollably" often describes the asynchronous thrashing, side-to-side head movements, or pelvic thrusting often seen in PNES.
- **Normal Interictal EEG:** While a single routine EEG is normal in ~50% of patients with true epilepsy, a completely normal EEG in a patient with frequent, severe motor events raises the suspicion of PNES.
- **Normal MRI:** This rules out structural causes such as tumors, mesial temporal sclerosis (scarring), or vascular malformations, which are common causes of new-onset seizures in adults.
- **Demographics/History:** The patient is a banker, a high-stress profession. PNES is classified as a functional neurological disorder and is often (though not always) associated with underlying stress or psychological factors (Conversion Disorder).

**2. Common Misconceptions in this Vignette**

You might hesitate to diagnose PNES because of the specific symptoms mentioned. However, clinical data clarifies these points:

- **Urinary Incontinence:** Historically, doctors were taught that incontinence only happens in "real" seizures. However, studies show that urinary incontinence occurs in **10–25% of PNES patients**. It is not a reliable exclusionary sign.
- **Injury/Falling:** Patients with PNES can and do sustain injuries during their episodes; the body's protective reflexes are not always preserved during a functional event.

**3. Differential Diagnosis**

While PNES is the likely answer given the negative workup, the following must be considered:

| **Diagnosis** | **Why it fits** | **Why it fits less** |
| --- | --- | --- |
| **Epilepsy (Low sensitivity EEG)** | Symptoms match; routine EEGs often miss deep focal epileptic spikes (e.g., Frontal Lobe Epilepsy). | MRI is normal; "shaking" description is atypical for organic seizure semiology. |
| **Convulsive Syncope** | Normal EEG/MRI; fainting can cause brief jerking due to hypoxia. | Incontinence is rare; "uncontrollable shaking" suggests a longer duration than the brief twitching of syncope. |
| **Alcohol Withdrawal** | "Banker" (social factors); withdrawal seizures are generalized tonic-clonic; MRI/EEG would be normal. | Vignette does not mention cessation of alcohol intake or other withdrawal signs (tremors, tachycardia). |

**4. Diagnostic "Gold Standard"**

You cannot diagnose PNES solely based on a normal routine EEG. The definitive diagnosis requires **Video-EEG Monitoring (vEEG)**.

- **The Goal:** To capture an episode on video while simultaneously recording brain waves.
- **The Result:** In PNES, the patient will be seen "shaking" and unresponsive on video, but the EEG trace will show normal background alpha rhythms (awake brain) rather than the chaotic high-amplitude spikes of a seizure.

**Next Step**

**Would you like me to explain the specific management and treatment therapies used for PNES, such as Cognitive Behavioral Therapy (CBT)?**

Based on the clinical presentation, the most likely diagnosis is **Idiopathic Generalized Epilepsy** (specifically appearing as generalized tonic-clonic seizures).

Here is a breakdown of why this diagnosis fits, despite the normal test results, and what the other considerations are.

**Reasoning for the Diagnosis**

**1. The Clinical Symptoms are Classic for Epilepsy** The combination of symptoms described is highly specific to a **Generalized Tonic-Clonic (Grand Mal) Seizure**:

- **Loss of Consciousness (LOC):** Immediate and total.
- **Uncontrollable Shaking:** Represents the clonic phase of the seizure.
- **Urinary Incontinence:** This is a high-specificity marker for epileptic seizures. While it can happen in other conditions (like syncope), it is statistically much more common in true epilepsy than in psychogenic non-epileptic seizures (PNES).
- **Post-ictal State:** While not explicitly detailed in the prompt, the "falling" and "shaking" usually lead to a period of confusion or sleepiness, which distinguishes it from syncope (where recovery is rapid).

**2. Why are the EEG and MRI Normal?** This is a common point of confusion, but it does not rule out epilepsy.

- **Normal MRI:** This is actually "good" news. It rules out structural causes like tumors, cavernous malformations, mesial temporal sclerosis, or neurocysticercosis (which might be suspected given the Texas location, but is ruled out by the scan). This points toward an **idiopathic** (genetic/metabolic/electrical) etiology rather than a structural one.
- **Normal EEG:** A standard EEG records only 20–30 minutes of brain activity. If the patient is not having a seizure at that exact moment, the EEG can be normal.
  - *Key Statistic:* Approximately **50% of patients** with true epilepsy will have a normal first interictal (between seizures) EEG.

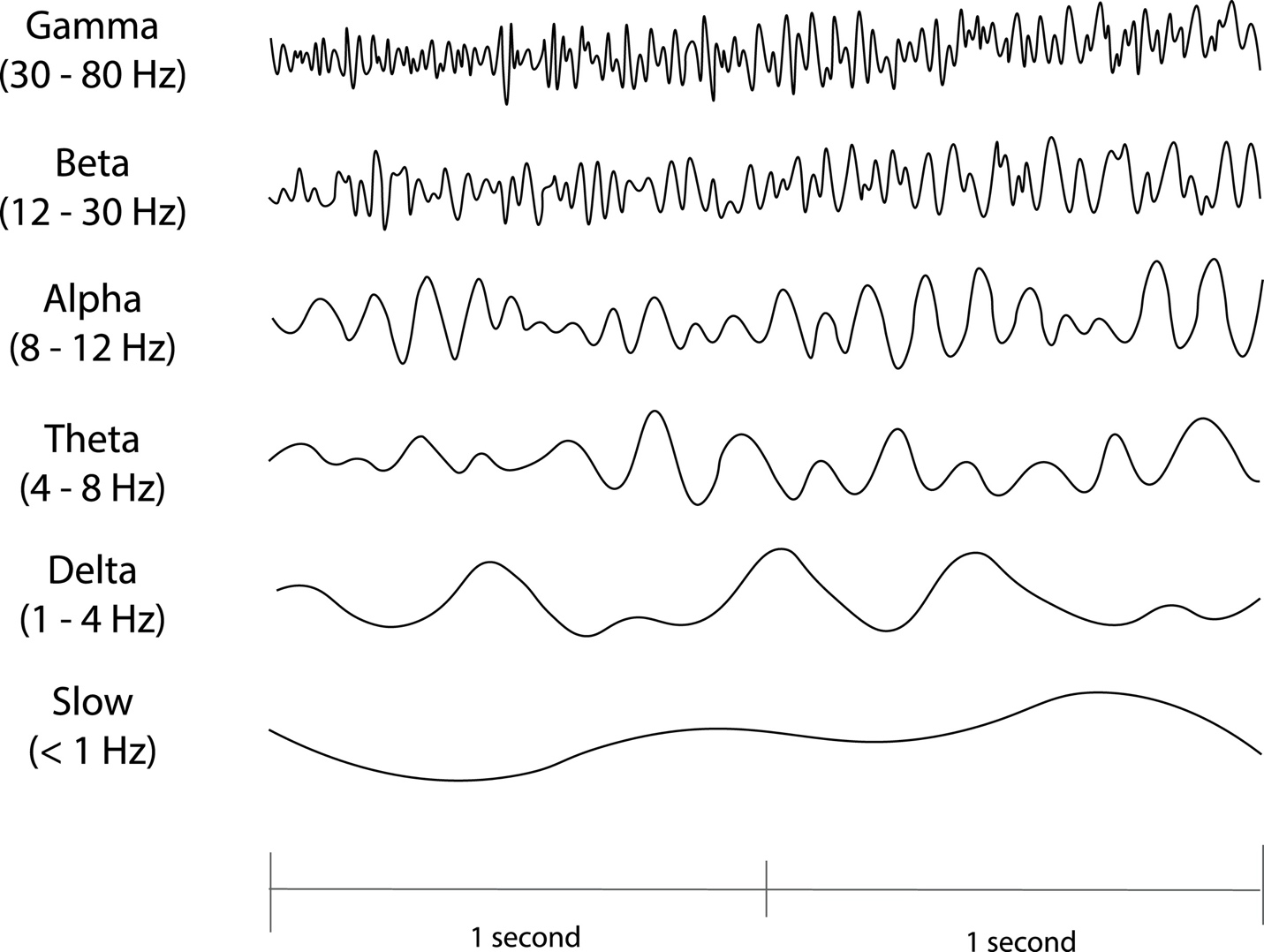

Shutterstock

**Differential Diagnoses (Ruled Out)**

| Condition | Why it is less likely |
| --- | --- |
| **Psychogenic Non-Epileptic Seizures (PNES)** | PNES (conversion disorder) is a strong consideration in patients with normal EEGs. However, **urinary incontinence** and significant injury from falling are much less common in PNES than in physiologic seizures. |
| **Convulsive Syncope** | Fainting (from heart issues or low blood pressure) can sometimes cause brief jerking. However, the jerking in syncope is usually brief/mild, and urinary incontinence is rare. |
| **Neurocysticercosis** | Being from Texas puts him in a higher risk zone for this parasitic infection. However, the **normal MRI** definitively rules this out, as the cysts would be visible. |
| **Alcohol Withdrawal** | Bankers are sometimes associated with high-stress, high-alcohol lifestyles in medical vignettes ("holiday heart," withdrawal). However, without a history of cessation of drinking 6–48 hours prior, this is speculative. |

Export to Sheets

**Summary**

The patient is suffering from **clinical seizures**. The "normal" workup implies that the cause is not a physical brain lesion (tumor/stroke) and that the electrical abnormality is not firing constantly.

**Next Step in Management:** To confirm the diagnosis, the standard next step is usually a **Sleep-Deprived EEG** or **Video-EEG Monitoring** to try and capture the event or interictal discharges that the first test missed.

Would you like me to explain the difference between a Tonic-Clonic and a Focal seizure?

Based on the clinical presentation and the specific combination of symptoms and test results, the most likely diagnosis is **Epilepsy (specifically Generalized Tonic-Clonic Seizures)**.

Despite the normal EEG and MRI, the clinical history of urinary incontinence and loss of consciousness strongly points toward an organic seizure disorder rather than a mimic.

**Key Reasons for This Diagnosis**

1. The Presence of "Hard Signs" (Urinary Incontinence)

While not present in every seizure, urinary incontinence is a statistically strong predictor of an organic seizure (epilepsy) as opposed to Psychogenic Non-Epileptic Seizures (PNES) or Syncope (fainting). In standard medical vignettes, incontinence and lateral tongue biting are often used to distinguish true seizures from other causes.

2. The Limitations of Routine EEGs

The fact that the EEG is normal is the "teaching point" of this scenario.

- **Interictal Period:** A routine EEG records brain activity between seizures (interictal).
- **Low Sensitivity:** A single routine EEG is normal in **30–50%** of patients with proven epilepsy.
- **Conclusion:** A normal EEG does not rule out epilepsy; it simply means the patient was not having a seizure *during*the test and did not show background abnormalities at that specific time.

3. The Normal MRI

The normal MRI rules out "secondary" causes of seizures, such as:

- Brain tumors
- Previous strokes
- Arteriovenous malformations (AVMs)
- Neurocysticercosis (relevant given the patient is from Texas, a border state, but ruled out by the scan)

This suggests the patient likely has **Idiopathic Generalized Epilepsy** (epilepsy with no structural cause, often genetic or unexplained).

**Differential Diagnosis (What else it could be)**

While Epilepsy is the primary diagnosis, a physician would consider the following based on the "Normal EEG" finding:

| **Condition** | **Why it fits** | **Why it is less likely** |
| --- | --- | --- |
| **Psychogenic Non-Epileptic Seizures (PNES)** | Normal EEG and MRI are hallmarks of PNES. High stress (Banker) can be a trigger. | Urinary incontinence is rare in PNES (though not impossible). The shaking in PNES is usually asynchronous (thrashing/pelvic thrusting) rather than rhythmic. |
| **Convulsive Syncope** | Fainting due to a heart issue or drop in blood pressure can cause brief jerking movements due to lack of oxygen to the brain. | "Shaking uncontrollably" usually implies a longer duration and intensity than the brief jerks of syncope. Incontinence is uncommon in syncope. |
| **Alcohol Withdrawal Seizure** | "Banker" is a demographic sometimes associated with high alcohol consumption. Withdrawal seizures show normal MRI and often normal interictal EEG. | This requires a history of heavy drinking and recent cessation, which is not provided in the prompt. |

**Recommended Next Step**

Because the routine EEG was normal but the clinical suspicion for epilepsy is high, the standard next step is **Video-EEG Monitoring** (often admitted to the hospital) or a **Sleep-Deprived EEG**. This allows doctors to capture the actual event or stress the brain to reveal the abnormalities missed by the first test.

Based on the clinical presentation and the specific details provided, the most likely diagnosis is **Epilepsy**, specifically **Generalized Tonic-Clonic Seizures** (formerly known as Grand Mal seizures).

Despite the normal EEG and MRI, the clinical history is the primary driver for this diagnosis.

**Reasoning**

**1. The Clinical Presentation is Classic** The patient describes the three distinct phases of a generalized tonic-clonic seizure:

- **Tonic Phase:** "Falling on the ground" and loss of consciousness (sudden loss of muscle tone or rigid stiffening).
- **Clonic Phase:** "Shaking uncontrollably" (rhythmic jerking of the limbs).
- **Autonomic Dysfunction:** "Urinary incontinence" is a common sign in generalized seizures and helps distinguish them from syncope (fainting) or psychogenic events.

**2. The "Normal" Workup (The Key Teaching Point)** This scenario is a classic medical board vignette designed to test a specific concept: **A normal interictal EEG does not rule out epilepsy.**

- **EEG:** A routine EEG measures brain waves *between* seizures (interictal). Approximately **50% of patients** with proven epilepsy have a completely normal first EEG. It often takes sleep-deprived EEGs or prolonged monitoring to find the abnormality.
- **MRI:** An MRI looks for structural causes (tumors, strokes, cavernomas, sclerosis). In **Idiopathic Generalized Epilepsy** (which often presents in young adulthood), the brain structure is anatomically normal.

**3. Why it is likely not other conditions**

- **Neurocysticercosis:** Being from Texas is a risk factor for this parasitic infection (pork tapeworm), which is a leading cause of adult-onset seizures. However, this is ruled out because the **MRI was normal** (cysts or calcifications would be visible).
- **Psychogenic Non-Epileptic Seizures (PNES):** These are stress-related events that mimic seizures. While the normal EEG supports this, the presence of urinary incontinence and significant injury (falling hard) makes organic epilepsy statistically more likely in a standard diagnostic context.
- **Syncope (Fainting):** While "convulsive syncope" exists (jerking after fainting), it usually resolves very quickly, and urinary incontinence is rare.

**Summary of Findings**

| Symptom/Test | Findings | Interpretation |
| --- | --- | --- |
| **Loss of Consciousness** | Yes | Indicates involvement of bilateral brain hemispheres or brainstem (RAS). |
| **Rhythmic Shaking** | Yes | Suggests motor cortex involvement (Clonic phase). |
| **Incontinence** | Yes | Strong indicator of a true epileptic event vs. a psychogenic event. |
| **Normal MRI** | Yes | Rules out tumor, stroke, or Neurocysticercosis. Points to Idiopathic Epilepsy. |
| **Normal EEG** | Yes | **Does not rule out epilepsy.** Simply means the patient was not seizing during the test. |

Export to Sheets

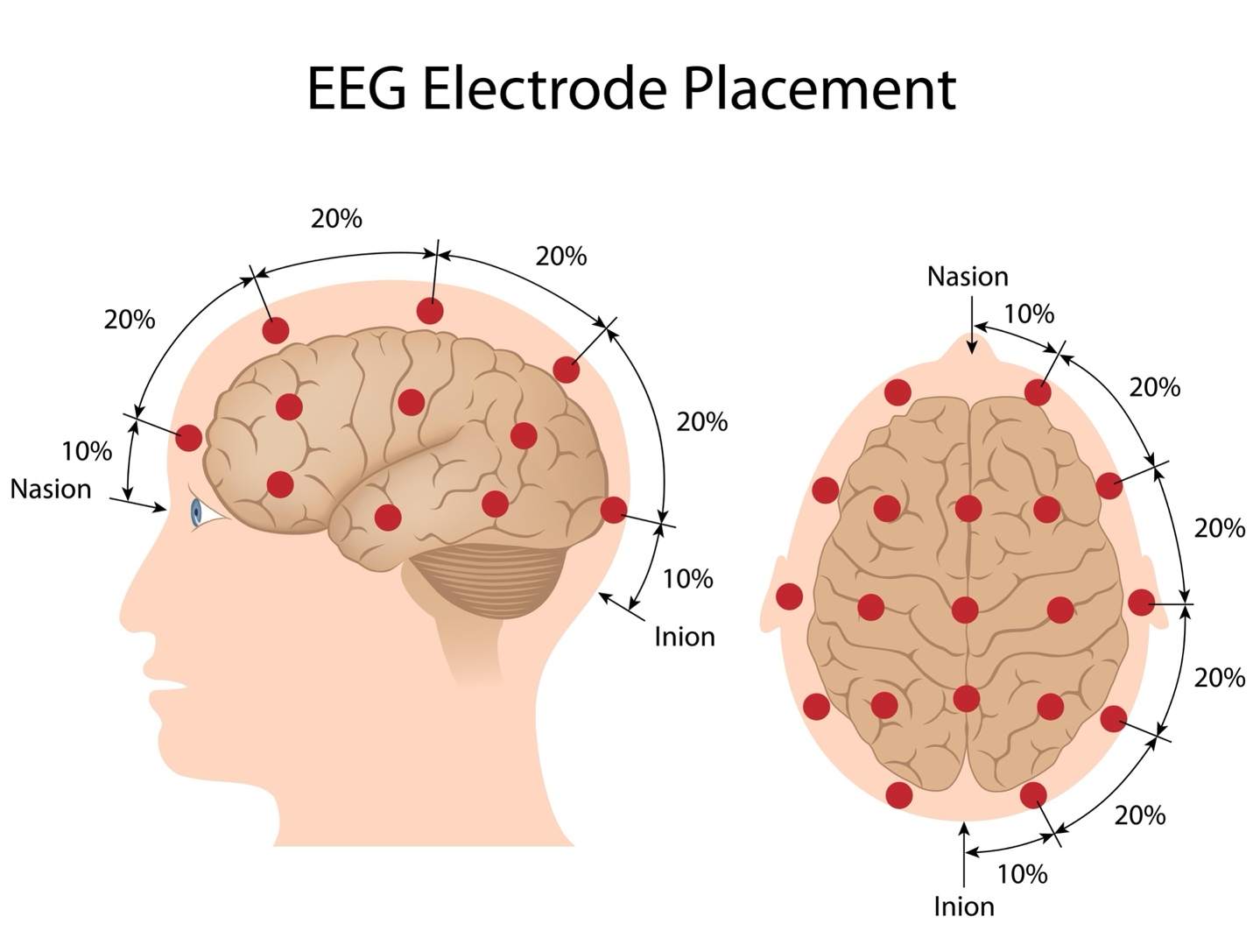

Shutterstock

**Next Steps**

Since the routine EEG was normal but the clinical suspicion is high, the standard next step is usually **Video-EEG Monitoring** (inpatient admission) to capture an actual spell, or a **Sleep-Deprived EEG** to increase the likelihood of provoking an abnormality.

**Would you like me to explain the specific differences between a Seizure and Syncope (fainting) to better differentiate the two?**

Based on the clinical presentation and the normal diagnostic results, the most likely diagnosis is **Psychogenic Non-Epileptic Seizures (PNES)**, also known as **Functional Seizures**.

While the symptoms mimic epilepsy, the combination of dramatic convulsive behavior with completely normal imaging and electrophysiology is the hallmark of this condition.

**Why this is the likely diagnosis**

1. **Normal EEG and MRI:**
   - **Epilepsy:** While a single interictal (between seizures) EEG can be normal in roughly 50% of patients with true epilepsy, the absence of any epileptiform discharges or slowing, combined with a normal MRI (ruling out tumors, mesial temporal sclerosis, or cavernomas), raises the suspicion for a non-epileptic etiology.
   - **PNES:** By definition, these patients have normal brain activity during the event. There is no abnormal electrical discharge in the brain.
2. **The Clinical Presentation:**
   - **"Shaking Uncontrollably":** In PNES, motor movements are often asynchronous (limbs moving out of phase), involve side-to-side head shaking, pelvic thrusting, or fluctuating intensity. This differs from the synchronous, rhythmic jerking of a generalized tonic-clonic seizure.
   - **Loss of Consciousness:** Patients with PNES often appear unconscious but may retain some awareness or have a "stuttering" return of consciousness, unlike the post-ictal confusion typical of epilepsy.
3. **The "Red Herrings" (Incontinence):**
   - Historically, urinary incontinence was taught as a hard sign of "true" epileptic seizures. However, modern research indicates that urinary incontinence occurs in a significant number of patients with PNES as well. Its presence does not rule out a functional disorder.

**Differential Diagnosis Considerations**

- **Epilepsy (Generalized Tonic-Clonic):** Still possible (as EEGs can be normal), but less likely without EEG correlates or structural abnormalities.
- **Convulsive Syncope:** Fainting due to a drop in blood pressure can cause jerking movements (myoclonus) due to brain hypoxia. However, these jerks are usually brief, and urinary incontinence is rare in syncope.
- **Malingering:** This is distinct from PNES. Malingering involves intentional faking for secondary gain (money, drugs). PNES is a subconscious **conversion disorder** (a somatic symptom disorder)—the patient is *not* faking and is truly suffering, but the cause is psychological/functional rather than electrical.

**The "Gold Standard" for Confirmation**

To confirm this diagnosis definitively, the patient requires **Video-EEG Monitoring (vEEG)**.

- The goal is to capture one of these "shaking" episodes on video while the patient is hooked up to the EEG.
- **Diagnosis Confirmation:** If the patient exhibits the shaking and unresponsiveness, but the EEG shows a normal background rhythm (alpha rhythm) without epileptic spikes, the diagnosis is confirmed as PNES.

**Would you like me to explain the recommended treatment approach for Functional Seizures?**

Based on the clinical vignette provided—a patient presenting with dramatic seizure-like episodes but a completely normal neurological workup (EEG and MRI)—the most likely diagnosis to consider is **Psychogenic Non-Epileptic Seizures (PNES)**, also known as **Functional Seizures**.

However, because a single routine EEG can be normal in patients with true epilepsy, this is a diagnosis of exclusion that requires careful differentiation.

**1. The Clinical Reasoning**

The diagnostic challenge here is distinguishing between organic pathology (Epilepsy/Syncope) and functional pathology (PNES).

| **Feature from Vignette** | **Interpretation** |
| --- | --- |
| **Falling & Shaking** | Suggests a generalized motor event. In true generalized tonic-clonic seizures, the EEG is almost always abnormal during the event, and interictal (between events) EEGs show abnormalities in ~50-70% of cases. |
| **Urinary Incontinence** | **The Distractor.** Historically, incontinence was thought to prove a seizure was "real" (epileptic). However, studies show incontinence occurs in **10–20% of patients with PNES**. It is not a definitive differentiator. |
| **Normal MRI** | Rules out structural causes like tumors, mesial temporal sclerosis, or neurocysticercosis (which is endemic in Texas/Mexico borders but would show calcifications or cysts on MRI). |
| **Normal EEG** | **The Key Clue.** While a single interictal EEG can be normal in epilepsy, the combination of recurrent, severe "generalized" convulsions with a completely normal electrical background strongly points toward a non-epileptic etiology. |
| **Demographics (Banker)** | High-stress professions can be a trigger for conversion disorders or stress-induced functional symptoms. |

**2. Differential Diagnosis**

While PNES is the leading suspect given the "normal" tests, other conditions must be weighed:

- **Epilepsy (False Negative):** A routine 30-minute EEG captures only a snapshot of brain activity. Approximately **50% of patients with true epilepsy** have a normal first EEG.
- **Convulsive Syncope:** A sudden drop in blood pressure (fainting) can cause hypoxic brain jerks that look like seizures. The patient is 32; if he has an undiagnosed arrhythmia (like Long QT syndrome), this could be fatal.
- **Alcohol Withdrawal:** Given the patient's age and profession (stereotypically associated with "work hard, play hard" cultures), alcohol withdrawal seizures are a possibility. These are generalized tonic-clonic seizures where the **MRI and interictal EEG are typically normal**.

**3. The Gold Standard for Diagnosis**

You cannot diagnose PNES solely based on a normal clinic visit. The diagnosis must be confirmed by capturing an event.

**Video-EEG Monitoring (vEEG)** is the required next step.

- The patient stays in the hospital for several days hooked up to a continuous EEG and video camera.
- **Diagnosis of PNES:** The patient has the "shaking" event, but the EEG shows normal waking alpha rhythm (no epileptic spikes).
- **Diagnosis of Epilepsy:** The EEG shows correlative epileptic discharges during the shaking.

**Summary**

The likely diagnosis is **Psychogenic Non-Epileptic Seizures (PNES)** due to the discordant findings (severe physical symptoms with normal electrical/structural brain health). However, **Alcohol Withdrawal** or **Epilepsy with a false-negative EEG** remain distinct possibilities until proven otherwise.

**Would you like me to explain the difference in treatment protocols for Epilepsy versus PNES?**

Based on the clinical vignette provided, the most likely diagnosis is **Idiopathic Generalized Epilepsy** (specifically Generalized Tonic-Clonic Seizures).

Even though the diagnostic tests (MRI and EEG) are normal, the clinical history is the primary driver for this diagnosis.

Here is a breakdown of the clinical reasoning, the significance of the normal test results, and the differential diagnoses that must be considered.

**1. Why the Diagnosis is Epilepsy**

Epilepsy is primarily a **clinical diagnosis**. The standard medical definition of epilepsy is having **two or more unprovoked seizures** occurring more than 24 hours apart. This patient has had three episodes, which meets the clinical criteria.

The symptoms described are classic for a **Generalized Tonic-Clonic Seizure (Grand Mal):**

- **Loss of Consciousness (LOC):** Sudden and complete.
- **Uncontrollable Shaking:** Represents the clonic (jerking) phase of the seizure.
- **Urinary Incontinence:** This is a specific sign that strongly points toward an organic "true" seizure rather than a fainting spell or psychological event. It indicates a total loss of sphincter control due to the magnitude of the cortical discharge.
- **Post-ictal State:** While not explicitly described in the text, the "falling" and "loss of consciousness" implies a period of recovery is needed.

**2. Interpreting the "Normal" Results**

It is a common misconception that a normal EEG and MRI rule out epilepsy.

- **Normal MRI:** This rules out *structural* causes (such as a brain tumor, stroke, cavernoma, or mesial temporal sclerosis). Because the brain structure is normal, the epilepsy is likely **Idiopathic** (arising from genetic or molecular causes) or **Cryptogenic** (obscure etiology) rather than "Symptomatic."
- **Normal EEG:** An EEG measures brain waves. If the patient is not having a seizure *during* the test (which is called an interictal EEG), the results can often be normal. **Approximately 50% of patients with epilepsy will have a normal first EEG.**

**3. Differential Diagnoses (What else could it be?)**

While Epilepsy is the leading diagnosis, a physician would consider these alternatives given the normal workup:

| **Condition** | **Description** | **Why it might fit** | **Why it might NOT fit** |
| --- | --- | --- | --- |
| **Psychogenic Non-Epileptic Seizures (PNES)** | Seizure-like events caused by psychological distress rather than electrical brain activity. | Normal EEG/MRI; High-stress job (Banker); Young adult. | **Urinary incontinence** is rare in PNES. The shaking in PNES is often asynchronous (side-to-side head movement), whereas epilepsy is usually synchronous. |
| **Convulsive Syncope** | Fainting due to lack of blood flow to the brain, followed by jerking movements. | Normal MRI/EEG; "Falling" suggests a drop in blood pressure. | The jerking in syncope is usually brief and myoclonic. "Uncontrollable shaking" and incontinence are much more indicative of a seizure. |
| **Alcohol Withdrawal Seizures** | Seizures triggered by cessation of alcohol intake ("Rum fits"). | Normal MRI/EEG between episodes; Banker occupation *could* imply lifestyle factors. | Requires a history of heavy drinking and timing related to cessation. |

**Summary**

The combination of **recurrent episodes** and **urinary incontinence** makes **Epilepsy** the most probable diagnosis. The normal MRI and EEG simply classify it as a non-lesional (no tumor/scar) epilepsy, likely genetic or metabolic in origin at the cellular level.

**Disclaimer:** *I am an AI, not a doctor. This analysis is for educational and informational purposes regarding a medical case study and does not constitute medical advice. If this is a real-life scenario, the patient should see a neurologist immediately.*

**Would you like me to explain the specific follow-up tests, such as Video-EEG monitoring, that would confirm this diagnosis?**

Based on the clinical presentation provided, the most likely diagnosis is **Epilepsy**, specifically manifesting as **Generalized Tonic-Clonic Seizures** (formerly known as Grand Mal seizures).

Despite the normal test results, the specific combination of symptoms strongly points toward an organic seizure disorder rather than a psychological or cardiovascular cause.

**Why this is the likely diagnosis**

The patient presents with the classic "tetrad" of symptoms associated with tonic-clonic seizures:

1. **Loss of Consciousness (LOC):** Sudden and complete.
2. **Tonic-Clonic Movements:** Described as "shaking uncontrollably" (rhythmic jerking).
3. **Falling:** Sudden loss of postural tone.
4. **Urinary Incontinence:** This is a high-value clinical clue. While not exclusive to epilepsy, urinary incontinence is a strong indicator of an organic event (a true seizure) and is statistically less common in psychogenic (stress-related) seizures or syncope (fainting).

**Understanding the "Normal" Tests**

It is a common misconception that a normal EEG and MRI rule out epilepsy. In this clinical vignette, the normal results actually help refine the type of epilepsy:

- **The Normal MRI:** This is good news. It rules out "Secondary Epilepsy" caused by structural issues, such as brain tumors, vascular malformations, strokes, or mesial temporal sclerosis.

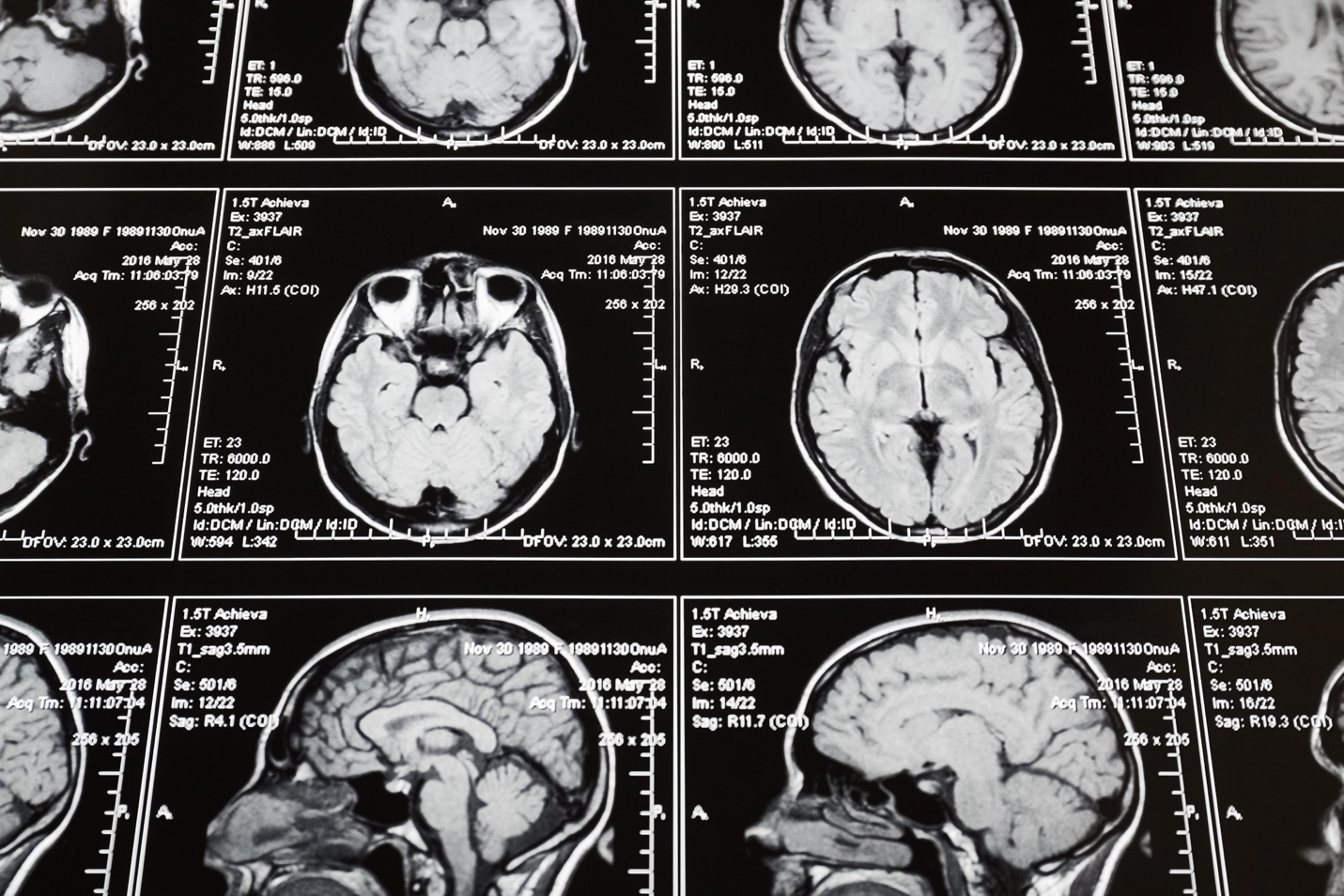

Shutterstock

- **The Normal EEG:** A standard EEG records brain waves for only 20–30 minutes. If the patient is not having a seizure *during* that specific 30-minute window, the brain waves often look normal. This is called an **interictal**(between seizures) EEG.
  - *Note:* Approximately **50% of patients** with true epilepsy will have a normal first EEG.

**Differential Diagnosis (What else it could be)**

While Epilepsy is the primary diagnosis, a physician would consider the following differentials:

| Condition | Why it fits | Why it is less likely |
| --- | --- | --- |
| **Psychogenic Non-Epileptic Seizures (PNES)** | Fits the "normal EEG" and "normal MRI" profile perfectly. | **Urinary incontinence** is much rarer in PNES than in true epilepsy. Also, PNES movements are often asynchronous (thrashing side-to-side) rather than rhythmic shaking. |
| **Convulsive Syncope** | Fainting (syncope) can sometimes cause brief jerking movements due to lack of blood flow to the brain. | Syncope recovery is usually very fast (seconds), whereas seizures usually have a "post-ictal" state (confusion/sleepiness) lasting minutes to hours. Incontinence is also less common in syncope. |
| **Metabolic Disturbance** | Can cause seizures (e.g., Alcohol withdrawal, hypoglycemia). | The patient has had **3 distinct episodes**, suggesting a chronic condition rather than a one-time metabolic crash. |

Export to Sheets

**Summary**

The diagnosis is **Epilepsy** (defined clinically as having two or more unprovoked seizures). The normal MRI suggests it is not caused by a tumor or injury (idiopathic/cryptogenic), and the normal EEG simply indicates the electrical activity is normal between events.

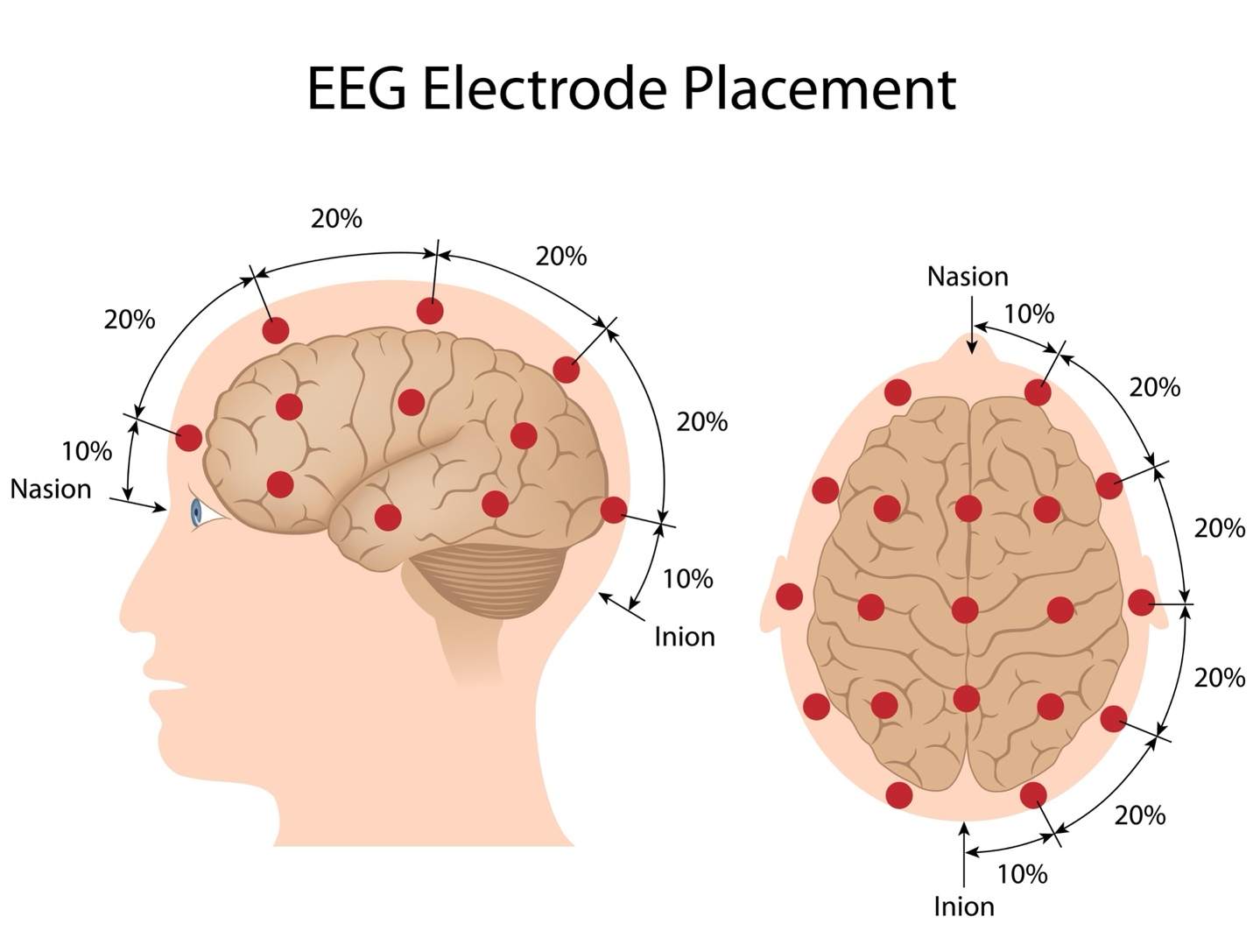

Shutterstock

**Next Step for the User**

To confirm this diagnosis definitively, a neurologist would likely order **Video-EEG Telemetry**. This involves admitting the patient to the hospital for 24–72 hours to capture an actual episode on camera while recording brain waves.

**Would you like me to explain the difference between a standard EEG and Video-EEG monitoring?**

Based on the clinical presentation and diagnostic results, the most likely diagnosis is **Epilepsy** (specifically **Generalized Tonic-Clonic Seizures**).

The presence of urinary incontinence and loss of consciousness strongly points toward an organic seizure disorder rather than a psychogenic cause, despite the normal imaging and EEG.

**Reasoning for the Diagnosis**

The "Normal EEG and MRI" in this vignette is a classic medical board "distractor" designed to test your understanding of seizure workups.

- **The Clinical Picture (Seizure vs. Imitators):**
  - **Urinary Incontinence:** This is a high-specificity indicator for organic epileptic seizures. It is rare in syncope (fainting) and uncommon in Psychogenic Non-Epileptic Seizures (PNES).
  - **"Shaking Uncontrollably" & Falling:** This describes a generalized tonic-clonic event.
  - **Recurrence:** The patient has had "3 episodes," meeting the clinical definition of epilepsy (two or more unprovoked seizures).
- **The Normal Diagnostic Tests:**
  - **Normal EEG:** A routine scalp EEG is **normal in approximately 50% of patients** with epilepsy during the interictal period (the time between seizures). A normal EEG does *not* rule out epilepsy.
  - **Normal MRI:** This rules out structural causes (tumors, strokes, cysts, vascular malformations). This suggests the epilepsy is likely **idiopathic** (genetic/chemical) or **cryptogenic** (hidden cause), rather than symptomatic of a lesion.

**Differential Diagnoses & Red Herrings**

The vignette includes specific demographic details ("Texas", "Banker") that act as red herrings or prompts for differential exclusion:

| Condition | Why it fits | Why it is unlikely (The "Rule Out") |
| --- | --- | --- |
| **Neurocysticercosis** | Common in **Texas** (and the Southwest) and a leading cause of adult-onset seizures. | The **MRI is normal**. Neurocysticercosis would typically show calcifications or cysts on brain imaging. |
| **Psychogenic Non-Epileptic Seizures (PNES)** | Fits the "Normal EEG/MRI" profile and the high-stress profession (**Banker**). | **Urinary incontinence** makes this statistically much less likely. Also, PNES movements are often described as asynchronous (thrashing) rather than the synchronous shaking of tonic-clonic seizures. |
| **Alcohol Withdrawal** | "Banker" may imply a high-stress/high-alcohol lifestyle ("liquid lunches"). Withdrawal seizures are tonic-clonic and have normal imaging. | Fits the "3 episodes" (if they were a cluster), but usually, a history of cessation is required. This remains a strong possibility if the history reveals heavy drinking. |
| **Convulsive Syncope** | Can cause falling and shaking (myoclonus) due to lack of blood flow. | **Incontinence** is rare in syncope. The shaking in syncope is usually brief and occurs *after* the fall, whereas seizures often cause the fall. |

Export to Sheets

**Recommended Next Steps**

The patient needs further evaluation to capture an event or increase the sensitivity of the EEG.

- **Sleep-deprived EEG:** Increases the chance of capturing epileptiform discharges.
- **Video-EEG Monitoring:** The gold standard to definitively confirm the diagnosis and rule out PNES.

**Would you like me to explain the specific differences in physical symptoms between a true epileptic seizure and a psychogenic seizure?**

[Seizures Review | Mnemonics And Proven Ways To Memorize for your exams!](https://www.youtube.com/watch?v=yaW0HhTDJX4) This video is relevant as it reviews the high-yield characteristics of different seizure types, including tonic-clonic seizures, which aligns with the symptoms described in your scenario.

[Seizures Review | Mnemonics And Proven Ways To Memorize for your exams! - YouTube](https://www.youtube.com/watch?v=yaW0HhTDJX4" \t "_blank)

[Cram The PANCE · 71K views](https://www.youtube.com/watch?v=yaW0HhTDJX4" \t "_blank)

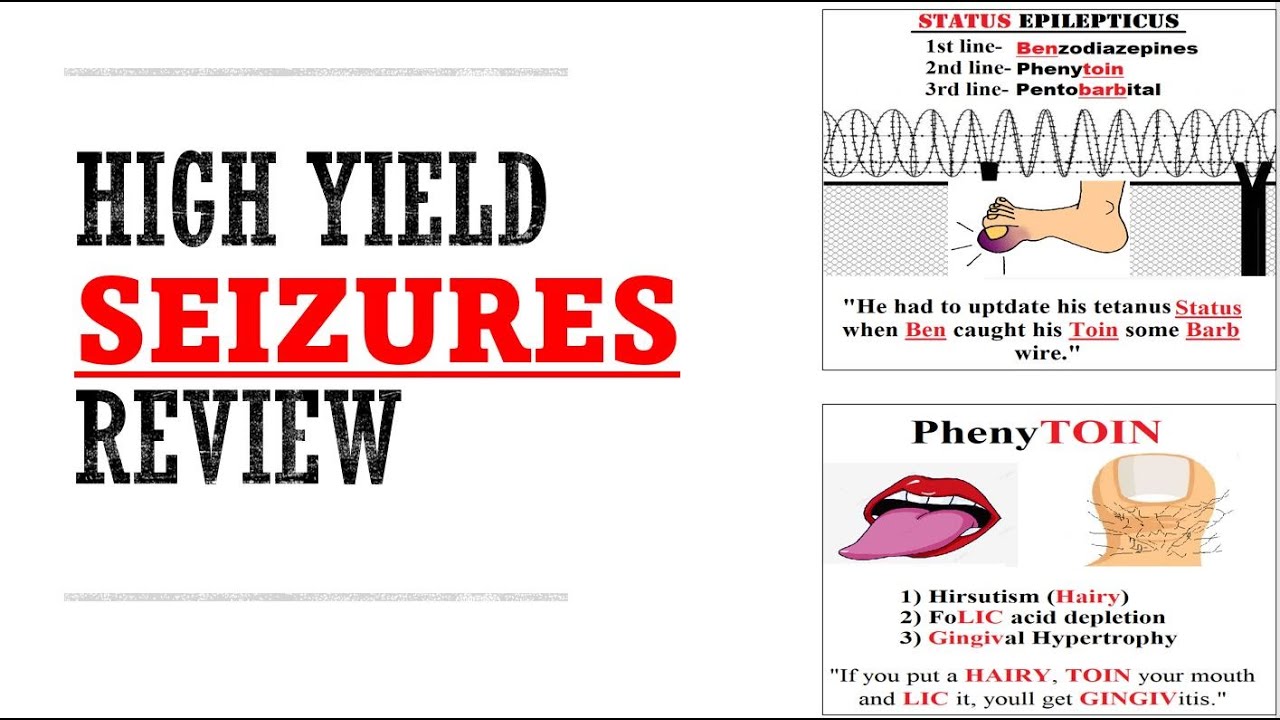
