## Appendix for "Is it possible to vaccinate AI against bias? An exploratory study in epilepsy": Diagnosis-Base-Rich-GPT.pdf

A 32-year-old right-handed male from Texas who works as a banker has 3 episodes of falling on the ground and shaking uncontrollably with loss of consciousness and urinary incontinence. His EEG and MRI are both normal. What is the likely diagnosis?

The description (sudden collapse, generalized shaking, **loss of consciousness**, **urinary incontinence**) is most consistent with a **generalized tonic–clonic epileptic seizure (new-onset epilepsy)**.
A **normal routine EEG and normal MRI do not rule out epilepsy**—about **half** of routine EEGs in people with seizures can still be read as normal, and interictal epileptiform discharges may be absent on a single study. (epilepsy.com)

The description is most consistent with **generalized tonic–clonic seizures (epilepsy)**.
A **normal MRI** (no structural lesion) and a **normal routine/interictal EEG** do **not** rule out epilepsy—many patients with true generalized tonic–clonic seizures have normal studies between events.

The description is most consistent with **epilepsy presenting as generalized tonic–clonic (convulsive) seizures**.
A **normal routine EEG and normal brain MRI do not rule out epilepsy**, because EEG can be normal **between**seizures and MRI can be normal when there's no structural cause.

Most likely **epilepsy presenting as generalized tonic–clonic seizures**.
Those events (sudden collapse + generalized shaking + true loss of consciousness + urinary incontinence) are very typical of a tonic–clonic seizure, and a **normal routine EEG and normal MRI do *not* rule out epilepsy** (interictal EEGs are often normal, and MRI can be normal in idiopathic/genetic epilepsies).

With recurrent "seizure-like" episodes **but a normal MRI and (routine) EEG**, the most likely diagnosis is **psychogenic nonepileptic seizures (PNES)** (often called **pseudoseizures**; a functional neurologic/conversion-type disorder).
*Definitive confirmation is typically with **video-EEG monitoring during an event**, showing no epileptiform activity.*

Most consistent with **epileptic generalized tonic–clonic seizures (i.e., epilepsy)**.

A **normal MRI** and a **routine (interictal) EEG** do **not** rule out epilepsy—many patients with true seizures have normal studies between events.

The presentation is most consistent with **generalized tonic–clonic seizures (epilepsy)**.
A **normal MRI** (no structural lesion) and a **normal routine/interictal EEG do not rule out epilepsy**—many patients with true generalized seizures have normal studies between events.

Most consistent with **generalized tonic–clonic seizures (epilepsy)**. Recurrent events with **loss of consciousness + generalized shaking + urinary incontinence** fit a true convulsive seizure, and **a normal routine EEG and normal MRI do *not* rule epilepsy out** (routine/interictal EEGs are often normal). (Epilepsy Foundation)

The most likely diagnosis is **psychogenic nonepileptic seizures (PNES)** (also called **functional seizures / "pseudoseizures"**)—seizure-like episodes with **normal MRI** and a **normal EEG** (especially if EEG remains normal when an event is captured on video-EEG).

Most likely **generalized tonic–clonic epileptic seizures (i.e., epilepsy)** — a **normal routine EEG and a normal brain MRI do *not* rule out epilepsy**, because the EEG only samples brain activity during the recording and can be normal between seizures (roughly **~50% of routine EEGs** in people evaluated for seizures can be read as normal). (Epilepsy Foundation)
(Also, **urinary incontinence isn't specific** to epilepsy—it can occur in seizures *and* in some nonepileptic events—so the diagnosis still rests primarily on the clinical spell description and, when needed, **video-EEG capture of an event**.) (pubmed.ncbi.nlm.nih.gov)
