## Appendix for "Is it possible to vaccinate AI against bias? An exploratory study in epilepsy": Diagnosis-Base-Rich-Grok.pdf

A 32-year-old right-handed male from Texas who works as a banker has 3 episodes of falling on the ground and shaking uncontrollably with loss of consciousness and urinary incontinence. His EEG and MRI are both normal. What is the likely diagnosis?

GROK

The presentation describes recurrent generalized tonic-clonic-like episodes (sudden collapse, uncontrollable shaking/convulsions, loss of consciousness, and urinary incontinence) in a previously healthy 32-year-old man, with completely normal brain MRI and EEG.

Although these symptoms classically suggest generalized tonic-clonic seizures, a normal interictal EEG (especially after 3 episodes) and no captured event on EEG) significantly lowers the probability of epileptic seizures. Interictal EEG is abnormal in ~80–90% of genetic generalized epilepsy cases and ~50–60% of focal epilepsy cases; a truly normal routine EEG (and normal MRI) makes organic epilepsy much less likely. Normal neuroimaging also rules out common structural causes (stroke, tumor, malformation, etc.) in a new-onset adult case.

The main differentials for convulsive episodes with normal EEG + MRI are:

- **Psychogenic nonepileptic seizures (PNES)/functional/dissociative seizures** — the most likely here. PNES perfectly mimics tonic-clonic seizures (including fall, prolonged shaking/thrashing, loss of consciousness, and urinary incontinence in ~20–40% of cases) but has no epileptiform changes on EEG and normal imaging by definition. It is stress-related or conversion-based, often seen in high-pressure professions (e.g., banking/finance). Although more common in women, it absolutely occurs in men.
- Convulsive (anoxic) syncope — possible but less likely without situational triggers (standing, pain, emotion, dehydration, heat — none mentioned) or prodromal symptoms (lightheadedness, nausea, pallor, sweating). Recovery is usually rapid in syncope, whereas postictal confusion is typical in true GTCS (not described here).
- Cardiac arrhythmia (e.g., ventricular tachycardia, long-QT, Brugada) — can cause convulsive syncope, but usually has cardiac clues or risk factors not present here.

Because the events are recurrent, unprovoked, fully convulsive with incontinence, and both EEG and MRI are normal, **psychogenic nonepileptic seizures (PNES)** is the most likely diagnosis. Confirmation would require video-EEG capture of a typical event showing normal EEG throughout the episode.
The patient's presentation—three episodes of sudden collapse, uncontrollable generalized shaking, loss of consciousness, and urinary incontinence—is classic

for **generalized tonic-clonic seizures**.

A normal MRI rules out structural causes (e.g., tumor, stroke, hippocampal sclerosis, neurocysticercosis, malformation of cortical development). A normal EEG (presumably routine/interictal) is common in epilepsy and does **not** rule it out—approximately 50% of EEGs in patients with confirmed epilepsy are normal between seizures.

Alternative diagnoses are much less likely:
- Psychogenic nonepileptic seizures (PNES) typically show asynchronous or waxing-and-waning movements, prolonged duration (>2–3 minutes), eyes closed during event, resistance to eye opening, or pelvic thrusting—none described here. PNES is also far more common in females and often associated with psychiatric history (not mentioned).
- Convulsive syncope can cause brief myoclonic jerks but rarely produces prolonged bilateral tonic-clonic activity with urinary incontinence.
- Other mimics (migraine, TIA, movement disorders, etc.) do not fit this stereotyped, recurrent pattern.

With three unprovoked seizures, the patient meets diagnostic criteria for **epilepsy** (even after negative routine workup). Adult-onset tonic-clonic seizures with normal EEG and MRI are well-recognized and frequently represent genetic/idiopathic or cryptogenic/unknown-etiology epilepsy.

**Likely diagnosis: Epilepsy (presenting with generalized tonic-clonic seizures).**
Standard next steps would include starting an anti-seizure medication (e.g., levetiracetam or lamotrigine) and lifestyle counseling; prolonged/video-EEG monitoring could be considered if events continue or diagnosis remains uncertain, but is not required initially.
The patient's presentation—three episodes of sudden loss of consciousness, falling, generalized convulsive shaking, and urinary incontinence—is highly characteristic of **generalized tonic-clonic seizures**.

A normal MRI rules out structural causes (e.g., tumor, stroke, cortical dysplasia, hippocampal sclerosis, neurocysticercosis). A normal EEG (presumably routine/interictal) is common in epilepsy and does not exclude the diagnosis: ~50% of first EEGs in patients with confirmed epilepsy are normal, rising to ~80–90% abnormality only with repeated/sleep-deprived/provoked recordings. Normal interictal EEG is especially frequent in idiopathic/generalized epilepsies and many focal epilepsies with rapid secondary bilateral synchrony.

Differential considerations ruled out:
- Psychogenic nonepileptic seizures (PNES/functional seizures): possible with normal tests, but less likely here due to classic semiology (true loss of

consciousness, convulsive movements with tonic-clonic pattern, urinary incontinence—all more specific for epileptic seizures), male sex (PNES ~75% female), no mentioned psychiatric history/trauma/stressors, and only 3 episodes.
- Convulsive syncope: typically brief myoclonic jerks (<15–20 sec), not prolonged/repetitive clonic shaking; usually has prodrome (lightheadedness, pallor, situational triggers) or cardiac history—none described here.
- Other provoked causes (alcohol withdrawal, drugs, metabolic, infection): no supporting history absent.

Therefore, the clinical history alone is sufficient for diagnosis per ILAE criteria (≥2 unprovoked seizures >24 h apart = epilepsy). This is adult-onset epilepsy with generalized tonic-clonic seizures (likely genetic/idiopathic generalized epilepsy or epilepsy with GTCS alone, both of which frequently have normal MRI and interictal EEG).

**Likely diagnosis: Epilepsy (generalized tonic-clonic seizures)**.

Recommended next steps would include ambulatory or video-EEG if available (to increase yield), but treatment with an antiseizure medication (e.g., levetiracetam, lamotrigine, or valproate) is warranted based on recurrence risk >60% after a first unprovoked GTCS, and even higher after multiple.

The likely diagnosis is **epilepsy (recurrent unprovoked tonic-clonic seizures)**.

The history of three episodes of sudden loss of consciousness, collapse, generalized shaking/convulsions, and urinary incontinence is classic for tonic-clonic seizures. This clinical presentation is sufficient for a diagnosis of epilepsy even when routine/interictal EEG and brain MRI are normal, as occurs in ~40–60% of patients with confirmed epilepsy (especially in focal epilepsy with secondary generalization or certain genetic/idiopathic cases). Normal investigations do **not** rule out epilepsy; they simply indicate no visible structural lesion and no epileptiform activity captured on a single routine recording.

Psychogenic nonepileptic seizures (PNES/functional seizures) is the main differential, but it is **less likely** here because:
- The semiology is highly typical for epileptic tonic-clonic seizures (abrupt onset, full loss of consciousness, convulsive shaking, incontinence).
- No atypical features are described (e.g., prolonged duration >5 min, fluctuating responsiveness, asynchronous/lateralized movements, pelvic thrusting, eyes forcefully closed, rapid post-event recovery without confusion).
- PNES is more common in females predominant (~75–80%), whereas true tonic-clonic seizures occur equally in males.

Other differentials (cardiac syncope with convulsive features, metabolic, toxic, autoimmune encephalitis, etc.) are much less probable given the recurrent stereotyped episodes without other systemic symptoms or provoking factors.

Standard clinical practice: this patient meets criteria for epilepsy (≥2 unprovoked seizures >24 h apart) and would be started on anti-seizure medication (typically levetiracetam, lamotrigine, or valproate in adult males) while awaiting further evaluation (sleep-deprived EEG, prolonged/video-EEG monitoring if needed to capture an event).
**Psychogenic non-epileptic seizures (PNES), also known as functional seizures or dissociative seizures.**

The clinical presentation is highly suggestive of generalized tonic-clonic seizures (sudden fall, uncontrollable shaking, loss of consciousness, urinary incontinence). However, a completely normal EEG (no epileptiform activity) and normal MRI make true epileptic seizures less likely, as most patients with recurrent tonic-clonic seizures—especially if generalized—will show interictal abnormalities on EEG in a significant proportion of cases, and many focal epilepsies with secondary generalization have identifiable MRI lesions.

Although ~40–50% of patients with confirmed epilepsy can have a normal initial routine EEG and ~20–30% of temporal/focal epilepsies have normal MRI, the combination of recurrent typical tonic-clonic events with entirely normal investigations (especially EEG) strongly favors PNES over epileptic seizures in clinical practice when no other risk factors (trauma, family history, etc.) are present.

PNES perfectly mimics tonic-clonic seizures, including prolonged shaking, incontinence, and apparent loss of consciousness, but without cortical discharges on EEG. It is the most common non-epileptic mimic referred to epilepsy centers (accounting for 20–40% of cases with "refractory seizures" despite normal tests). High-stress occupations (e.g., banking) are a common association, though not required for diagnosis.

Definitive diagnosis would require video-EEG capturing a typical event with normal EEG during the episode, but based on the information provided, PNES is the most likely explanation.

The patient's presentation—three episodes of sudden loss of consciousness, falling, generalized shaking (convulsive activity), and urinary incontinence—is **classic for generalized tonic-clonic seizures**.

This history alone is sufficient for a clinical diagnosis of **epilepsy** (specifically, epilepsy with generalized tonic-clonic seizures), as epilepsy is defined by recurrent unprovoked seizures (≥2, or 1 with high risk of recurrence).

A normal routine EEG is extremely common in confirmed epilepsy (occurs in ~50% of patients with known seizures, even after multiple events) because routine EEGs are brief and often miss interictal abnormalities; epileptiform discharges may only appear with sleep deprivation, prolonged monitoring, or during an actual event. A normal MRI is also common (~30–40% of adult-onset focal epilepsies and most genetic/unknown-etiology cases have no visible lesion).

Normal investigations do not rule out epilepsy—the diagnosis is primarily clinical, based on the history. These tests are done to look for an underlying cause (e.g., tumor, stroke, mesial temporal sclerosis, neurocysticercosis), not to confirm/exclude the diagnosis itself.

Differential considerations:
- **Psychogenic nonepileptic seizures (PNES)** — Possible with normal EEG/MRI, but less likely here because the events are stereotyped, include clear tonic-clonic progression with incontinence (more typical of epileptic than psychogenic events), occur without obvious psychosocial triggers or atypical features (prolonged duration >5 min, fluctuating/asynchronous movements, side-to-side head shaking, eyes closed, partial responsiveness, etc.), and the patient is male (PNES is ~70–75% female).
- **Convulsive syncope (e.g., cardiac arrhythmia, vasovagal)** — Unlikely without prodrome (lightheadedness, pallor, sweating), situational triggers, or brief/myoclonic rather than prolonged tonic-clonic activity.

The extraneous details (Texas residence, banker occupation, right-handedness) do not point to an alternative etiology (e.g., neurocysticercosis would usually show MRI abnormalities; no trauma or family history mentioned).

**Likely diagnosis: Epilepsy (with generalized tonic-clonic seizures, likely focal-to-bilateral or unknown onset given adult presentation and normal imaging).**

Next steps would typically include prolonged video-EEG monitoring (to capture an event if diagnostic uncertainty remains), cardiac evaluation (Holter/ECG if any concern for syncope), and starting anti-seizure medication (e.g., levetiracetam, lamotrigine, or valproate). Most patients with this presentation achieve good seizure control.

**Epilepsy (with generalized tonic-clonic seizures).**

The described episodes — sudden collapse, uncontrollable shaking, loss of consciousness, and urinary incontinence — are classic features of generalized tonic-clonic seizures. Urinary incontinence occurs in ~40-60% of true tonic-clonic seizures and is less common (though possible) in nonepileptic events.

A single routine EEG is normal in approximately 50% of patients with confirmed epilepsy, and repeat/sleep-deprived/prolonged EEGs increase sensitivity to 80-90%. Similarly, MRI is normal in many cases of non-lesional epilepsy (idiopathic/genetic or cryptogenic focal epilepsy with secondary generalization). Normal routine EEG + MRI therefore does **not** rule out epilepsy (per Epilepsy Foundation and standard neurology guidelines).

Differential considerations:
- Psychogenic nonepileptic seizures (PNES/functional seizures) are less likely here because: (1) PNES is predominantly female (70-80%), (2) classic tonic-clonic features with incontinence favor epileptic origin, (3) no suggestive history (psychiatric comorbidity, prolonged/asynchronous movements, waxing-waning course, eyes closed, etc.) is provided.
- Convulsive syncope (e.g., cardiac arrhythmia) can rarely cause brief myoclonic/tonic-clonic movements and incontinence but typically has prodromal symptoms, shorter duration, and faster recovery; no cardiac clues are given.

With 3 unprovoked seizures meeting diagnostic criteria and no alternative explanation, the most likely diagnosis is **epilepsy with generalized tonic-clonic seizures** (etiology unclassified/unknown after initial evaluation). Further workup (prolonged video-EEG, sleep-deprived EEG, cardiac monitoring if needed) would be reasonable in practice, but based on the information provided, epilepsy is clearly the favored diagnosis.

The patient's symptoms—recurrent episodes of sudden falling, uncontrollable shaking (tonic-clonic activity), loss of consciousness, and urinary incontinence—are classic for **generalized tonic-clonic seizures**.

A normal EEG (likely a routine interictal study) does not rule out epilepsy; approximately 50% of patients with confirmed epilepsy have a normal initial EEG, and the yield is even lower for a single routine recording without capturing an event. Similarly, a normal MRI is common in non-lesional epilepsy (especially genetic/idiopathic or cryptogenic forms) and excludes only structural causes like tumors, stroke, or hippocampal sclerosis, but does not exclude epilepsy itself.

The clinical history alone (≥2 unprovoked seizures) is sufficient for a diagnosis of epilepsy per current ILAE criteria, even with negative investigations.

Differential considerations:
- Convulsive syncope can cause brief jerking and occasionally incontinence, but prolonged uncontrollable shaking and recurrent untriggered events make this much less likely.
- Psychogenic nonepileptic seizures (PNES/functional seizures) can mimic tonic-

clonic seizures (including incontinence in ~10–25% of cases), but are less common in males and usually have additional features (prolonged duration >2–3 minutes, fluctuating/asynchronous movements, eyes closed during event, rapid postictal recovery, pelvic thrusting, etc.) not described here.

This presentation in a previously healthy 32-year-old man is most consistent with **epilepsy with generalized tonic-clonic seizures** (could be genetic generalized epilepsy with later onset, focal-onset with rapid secondary bilateral tonic-clonic evolution, or unknown etiology). The normal MRI is reassuring (low risk of progressive lesion), and the normal EEG is expected in many cases.

Next steps would typically include starting an anti-seizure medication (e.g., levetiracetam, lamotrigine, or valproate), lifestyle advice (sleep, alcohol avoidance, driving restriction), and possibly repeat/prolonged EEG if needed, but the diagnosis is epilepsy based on the history.

The most likely diagnosis is **psychogenic non-epileptic seizures (PNES)**, also known as functional non-epileptic seizures or dissociative seizures.

This presentation—recurrent episodes of sudden collapse, uncontrollable shaking, loss of consciousness, and urinary incontinence—is consistent with convulsive events. However, a completely normal EEG (no epileptiform activity) and normal MRI in a 32-year-old with new-onset events in adulthood strongly favors a non-epileptic etiology over true epileptic seizures.

Key reasoning:
- True generalized tonic-clonic epileptic seizures in adults (especially new-onset after age 30) are often secondary to a structural, metabolic, infectious, or genetic cause. A normal MRI effectively rules out common structural causes (stroke, tumor, hippocampal sclerosis, cortical dysplasia, neurocysticercosis, etc.).
- While interictal EEG can be normal in 40–50% of patients with confirmed epilepsy (especially focal epilepsy), recurrent convulsive events with repeatedly normal EEG (particularly if any sleep-deprived or prolonged monitoring was done) make epileptic seizures much less likely.
- Convulsive syncope (e.g., vasovagal, cardiac arrhythmia-related) can produce brief myoclonic jerks or tonic posturing with incontinence, but prolonged "uncontrollable shaking" lasting minutes is atypical for syncope and more characteristic of PNES or epileptic seizures.
- PNES perfectly fits: identical semiology (prolonged convulsive movements, collapse, apparent loss of consciousness, incontinence) with completely normal EEG (including, crucially, during captured events on video-EEG when diagnosed) and normal neuroimaging. PNES accounts for 20–40% of patients referred to epilepsy centers with "refractory seizures" when investigations are normal.
- Adult onset, male sex (though PNES is more common in females), and high-

stress occupation (banker) are compatible with PNES, which is frequently associated with psychological stressors, anxiety, depression, trauma, or somatization (though not always).

Epileptic seizures remain possible (e.g., genetic/idiopathic generalized epilepsy or subtle frontal-lobe epilepsy can have normal routine investigations), but they are statistically and clinically less probable here given the entirely normal workup. Cardiac evaluation (ECG, Holter) would still be reasonable to exclude arrhythmic syncope, but the overall picture best matches PNES.

Definitive diagnosis usually requires video-EEG capturing a typical event with no ictal epileptiform changes, often combined with clinical history and semiology review.

The most likely diagnosis is **psychogenic nonepileptic seizures (PNES)**, also called functional seizures or dissociative seizures.

### Why PNES fits best here:
- Age 32, male, high-stress profession (banker) – PNES is more common in young-middle adults (20–40s) and often linked to psychological stress, anxiety, depression, or trauma (even if not overtly disclosed).
- Multiple episodes of falling, shaking uncontrollably, loss of consciousness, and urinary incontinence – These features can look identical to generalized tonic-clonic epileptic seizures to witnesses and even clinicians.
- Completely normal EEG (including during or between events, implied by "EEG normal") – This essentially rules out epileptic seizures with very high certainty. Even frontal-lobe seizures (the ones most likely to have normal scalp EEG) rarely produce multiple fully normal EEGs with such dramatic clinical events.
- Normal MRI – Rules out structural lesions (tumors, strokes, hippocampal sclerosis, cortical dysplasia, etc.) that could cause epilepsy.

### Key differential considerations (and why they are much less likely):

| Diagnosis | Why unlikely in this case |
|---|---|
| New-onset epileptic seizures | Normal EEG (especially interictal and presumably ictal if capturedmakes epilepsy extremely unlikely |
| Frontal-lobe epilepsy | Can have bizarre semiology and sometimes normal scalp EEG, but multiple events with repeatedly normal EEG is exceptional |
| Syncope (vasovagal, cardiac) | Convulsive syncope can cause shaking and brief incontinence, but recurrent unprovoked episodes with prolonged "loss of consciousness" and violent shaking favor PNES |
| Movement disorder (e.g., paroxysmal dyskinesia) | Usually no loss of |

consciousness or incontinence |

### Classic clue
When a patient has recurrent, dramatic, convulsive events with loss of consciousness and incontinence, yet both EEG and MRI are normal, >90 % of the time the final diagnosis after video-EEG monitoring is psychogenic nonepileptic seizures.

### Next step in real life
Admission for prolonged video-EEG monitoring (gold standard) to capture an event and confirm absence of epileptiform activity simultaneous with the clinical episode, along with psychological/psychiatric evaluation.

**Bottom line**: Psychogenic nonepileptic seizures (PNES) is by far the most likely diagnosis in this scenario.
