## Appendix for "Is it possible to vaccinate AI against bias? An exploratory study in epilepsy": Diagnosis-Base-Rich-Kimi.pdf

A 32-year-old right-handed male from Texas who works as a banker has 3 episodes of falling on the ground and shaking uncontrollably with loss of consciousness and urinary incontinence. His EEG and MRI are both normal. What is the likely diagnosis?

KIMI

Based on the clinical presentation and normal investigations, the **most likely diagnosis is vasovagal syncope** (a form of reflex syncope), with psychogenic non-epileptic seizures (PNES) as the primary differential diagnosis. True epileptic seizures are less likely but cannot be completely excluded.

### Key Diagnostic Considerations

#### Why Vasovagal Syncope is Most Likely

The presentation of falling, shaking, loss of consciousness, and urinary incontinence can all occur in vasovagal syncope:

- **Motor activity**: Stiffening and tonic-clonic movements occur in **at least 50% of syncopal episodes**, typically lasting seconds and often described as "shaking"
- **Urinary incontinence**: Is **common in syncope** and has **no discriminatory value** in differentiating syncope from seizures (sensitivity 38%, specificity 57%)
- **Normal investigations**: Interictal EEG is typically normal in syncope and **cannot exclude the diagnosis**
- **Epidemiology**: Syncope is **commonly misdiagnosed as epilepsy**, with misdiagnosis rates of 4.6-30%

#### Differential Diagnosis

**1. Psychogenic Non-Epileptic Seizures (PNES)**
- Also strongly considered given normal EEG and MRI
- Can present with identical motor features, loss of consciousness, and incontinence
- Definitive diagnosis requires **video-EEG monitoring** showing typical episodes without EEG correlation
- More common in women but can occur in men, often associated with psychological stress

**2. Epilepsy (Less Likely)**
- While focal or generalized epileptic seizures can present this way, **normal EEG and MRI make this less probable**

- Interictal EEG may be normal in 30-55% of epilepsy patients initially, but serial studies detect abnormalities in 80-90%
- Postictal confusion typically lasts >10 minutes in epilepsy versus brief recovery in syncope

#### Critical Next Steps

1. **Detailed history**: Identify triggers (pain, emotional stress, prolonged standing, dehydration), prodromal symptoms (lightheadedness, nausea, visual changes), and post-event recovery time
2. **Cardiac evaluation**: ECG and possibly tilt-table testing to confirm syncope and rule out cardiac causes
3. **Video-EEG monitoring**: The gold standard to differentiate between epileptic seizures, PNES, and syncope
4. **Witness accounts**: Critical for distinguishing episode characteristics

**Conclusion**: Given the normal neuroimaging and EEG, coupled with the high prevalence of syncope mimicking seizures, **vasovagal syncope** is the most probable diagnosis, but **PNES must be excluded** through video-EEG monitoring before establishing a definitive diagnosis.

Based on the clinical presentation of 3 episodes with loss of consciousness, convulsive shaking, and urinary incontinence—but with normal EEG and MRI—the most likely diagnosis is **Psychogenic Nonepileptic Seizures (PNES)**.

#### Key diagnostic reasoning:

**Why PNES is most likely:**
- **Normal EEG**: The lack of epileptiform activity is highly suggestive of PNES. While a single normal EEG doesn't completely rule out epilepsy (since interictal studies can be normal), the combination with normal MRI makes epilepsy less likely.
- **Epidemiology**: PNES affects 20-40% of patients referred to epilepsy centers for refractory seizures, making it a common cause of seizure-like episodes.
- **Clinical features**: The described episodes (falling, convulsive movements, loss of consciousness, urinary incontinence) can all occur in PNES, which mimics epileptic seizures closely.

**Important considerations:**

1. **Urinary incontinence is not discriminatory**: Research shows urinary incontinence occurs in both epileptic seizures and non-epileptic events (syncope, PNES) and cannot reliably distinguish between them.

2. **Epilepsy is still possible**: Normal interictal EEG and MRI do not completely exclude epilepsy, especially if seizures are infrequent. Up to 50% of patients with epilepsy may have normal routine EEGs. However, in a test question context, normal studies strongly point toward PNES.

3. **Convulsive syncope**: Brief convulsive movements can occur with syncope, but these episodes are typically shorter and have clear triggers (pain, prolonged standing, dehydration). The occupational context (banker) doesn't provide typical syncope triggers.

**Recommended next steps:**
- **Video-EEG monitoring**: The gold standard for definitive diagnosis, capturing an episode to confirm absence of epileptiform activity during the event
- **Psychological evaluation**: To identify underlying stress, trauma, or psychiatric comorbidities
- **Multidisciplinary approach**: Neurology and psychology collaboration for proper diagnosis and treatment

The diagnosis requires careful, empathetic communication as PNES represents genuine, involuntary events—not "faking"—and patients often benefit from cognitive behavioral therapy and stress management.

Based on the clinical presentation and normal investigations, the most likely diagnosis is **Epilepsy with Generalized Tonic-Clonic (Grand Mal) Seizures**, though **Psychogenic Non-Epileptic Seizures (PNES)** must be considered as an important differential diagnosis.

#### Primary Diagnosis: Generalized Tonic-Clonic Seizures

The patient's symptoms—falling, uncontrollable shaking, loss of consciousness, and urinary incontinence—are classic for generalized tonic-clonic seizures. Several key points support this diagnosis:

1. **Typical Semiology**: The combination of convulsive activity, loss of consciousness, and incontinence is characteristic of tonic-clonic seizures, which involve violent muscle contractions followed by rhythmic jerking.

2. **Normal EEG Doesn't Exclude Epilepsy**: Approximately 50% of patients with epilepsy have a normal first interictal EEG. Epileptiform abnormalities may only be captured during a seizure or with repeated studies.

3. **Normal MRI is Common**: Many epilepsy patients, particularly those with genetic or idiopathic generalized epilepsy, have normal brain MRI scans.

4. **Urinary Incontinence Mechanism**: Tonic-clonic seizures cause pelvic muscle tension and increased intra-abdominal pressure, leading to bladder leakage in up to 39% of epilepsy patients.

#### Important Differential: Psychogenic Non-Epileptic Seizures (PNES)

Given the normal investigations, PNES warrants strong consideration:

- **Presentation**: PNES can mimic tonic-clonic seizures with convulsions, loss of consciousness, and urinary incontinence, but shows **normal EEG findings**.
- **Risk Factors**: The patient's high-stress occupation as a banker aligns with PNES etiology, which is often triggered by mental stress, emotional trauma, or psychological conditions.
- **Demographics**: While PNES is more common in women, it can occur in men with significant stressors.

#### Distinguishing Features to Assess

Further evaluation should focus on:
- **Postictal state**: True seizures typically cause prolonged confusion; PNES often lacks this
- **Episode duration**: PNES episodes often last longer (>5 minutes) with waxing/waning intensity
- **Injury pattern**: Tongue biting (especially lateral) and other injuries more common in true seizures
- **Video-EEG monitoring**: Gold standard to capture events and differentiate between epileptic and psychogenic causes

#### Conclusion

While the classic semiology points to **generalized tonic-clonic epilepsy**, the normal EEG and MRI in the context of a high-stress occupation make **PNES** a critical differential that requires video-EEG monitoring for definitive diagnosis.

Based on the clinical presentation and normal diagnostic studies, the most likely diagnosis is **non-epileptic seizures (NES)**, specifically psychogenic non-epileptic seizures (PNES).

#### Key Diagnostic Considerations:

**1. Non-Epileptic Seizures (Most Likely)**
The combination of convulsive activity ("shaking uncontrollably"), loss of consciousness, urinary incontinence, and **normal EEG/MRI** strongly suggests

NES. According to Cedars-Sinai, NES can present with "convulsions, falling down, loss of consciousness, loss of bladder control" while showing **no unusual electrical activity on EEG** and typically normal MRI findings. NES is most often caused by mental stress or psychological factors rather than abnormal brain electrical activity.

**2. Why Not Atonic Seizures?**
While the "falling on the ground" could suggest atonic seizures, the **"shaking uncontrollably"** description doesn't fit. Atonic seizures involve sudden **loss of muscle tone** without prominent shaking, typically last <15 seconds, and the person often remains conscious. The semiology described is more consistent with tonic-clonic activity.

**3. Important Exclusions Before Confirming NES:**
- **Convulsive syncope**: Can mimic seizures with shaking and urinary incontinence; cardiac evaluation is essential
- **Interictal EEG limitations**: A single routine EEG may miss epileptiform activity; video-EEG monitoring during an event is the gold standard
- **Cardiac arrhythmia**: As highlighted in a reported case, urinary incontinence with loss of consciousness can rarely be cardiac-related

#### Recommended Next Steps:
1. **Video-EEG monitoring** to capture an episode and confirm absence of epileptiform activity
2. **Psychiatric/psychological evaluation** to identify underlying stressors or psychological factors
3. **Cardiac workup** (ECG, Holter monitor) to exclude arrhythmia
4. **Detailed witness interviews** to better characterize the episodes

**Final Answer**: Psychogenic non-epileptic seizures is the most likely diagnosis, but this remains a diagnosis of exclusion requiring comprehensive evaluation.

Based on the clinical presentation described, the **most likely diagnosis is epilepsy with generalized tonic-clonic seizures (GTCS, also known as grand mal seizures)**.

### Rationale for Diagnosis

The key features strongly suggest epileptic seizures:
- **Falling with convulsive shaking**: Classic motor manifestation of GTCS
- **Loss of consciousness**: Invariable in GTCS
- **Urinary incontinence**: Common in GTCS due to detrusor muscle contraction and sphincter relaxation during the seizure
- **Multiple episodes**: Recurrent unprovoked events meeting epilepsy criteria

### Critical Point: Normal EEG and MRI Do Not Exclude Epilepsy

**Normal EEG:**
- Interictal EEG (recorded between seizures) is normal in many patients with epilepsy
- Epileptiform abnormalities may be absent in routine EEGs, especially if seizures are infrequent
- Studies show that "a normal EEG does not exclude epilepsy"
- Many patients require repeat EEGs, sleep-deprived EEGs, or prolonged video-EEG monitoring to capture abnormalities

**Normal MRI:**
- Many epilepsy syndromes show no structural abnormalities on MRI
- MRI negativity is particularly common in genetic or idiopathic epilepsies
- The brain may appear structurally normal despite abnormal electrical activity

### Differential Diagnoses to Consider

1. **Convulsive Syncope**:
   - Can present with brief myoclonic jerks and incontinence
   - Usually has identifiable triggers (prolonged standing, emotional stress, dehydration)
   - Episode duration is typically shorter (<30 seconds) and recovery is rapid
   - The description "shaking uncontrollably" suggests more prolonged convulsive activity than typical syncope

2. **Psychogenic Nonepileptic Seizures (PNES)**:
   - Can mimic epileptic seizures but often have atypical features
   - Incontinence is less common and usually not accompanied by true convulsive activity

### Recommended Next Steps

1. **Video-EEG monitoring**: Capturing an event is the gold standard for definitive diagnosis
2. **Cardiac evaluation**: ECG and Holter monitoring to exclude cardiac syncope, especially given reports that "urinary incontinence without convulsions might be a red flag for cardiogenic syncope"
3. **Laboratory tests**: Basic metabolic panel, toxicology screen
4. **History review**: Identify potential triggers, prodromal symptoms, and postictal features

### Prognosis

With appropriate antiseizure medication, more than half of patients achieve seizure freedom after their first medication trial . The normal EEG and MRI in a young adult suggest idiopathic epilepsy, which often has a good prognosis.

**Final Answer**: The presentation is most consistent with **epilepsy with generalized tonic-clonic seizures**, despite normal EEG and MRI, as these investigations can be completely normal in many patients with epilepsy.

Based on the clinical presentation described, the **most likely diagnosis is generalized tonic-clonic seizures (GTCS) from epilepsy**, despite the normal EEG and MRI. However, psychogenic non-epileptic seizures (PNES) and convulsive syncope must also be considered in the differential diagnosis.

### Key Diagnostic Considerations

**Why GTCS/Epilepsy is most likely:**
- The clinical semiology is classic: **uncontrollable shaking with loss of consciousness, falling, and urinary incontinence** are hallmark features of generalized tonic-clonic seizures
- While EEG and MRI are normal, this **does NOT exclude epilepsy**: approximately 50% of interictal EEGs in patients with seizures are normal because they only record brain activity between seizures
- You can have epilepsy with completely normal neuroimaging (MRI) and normal interictal EEG

**Important caveats from the literature:**

1. **Normal EEG is not exclusionary**: The EEG only captures activity during the test period. If the patient wasn't having a seizure at that moment, abnormal activity may not be recorded . Even patients with frequent seizures can have normal routine EEGs.

2. **Urinary incontinence is non-specific**: A systematic review found that urinary incontinence has **poor discriminatory value** between epileptic seizures and non-epileptic events, with only 38% sensitivity and 57% specificity . It occurs in both epilepsy, syncope, and PNES.

3. **Other possibilities to consider**:
   - **Psychogenic Non-Epileptic Seizures (PNES)**: Can mimic GTCS with motor activity and incontinence. Normal EEG is typical. More common in women but can affect men. Requires **video-EEG monitoring** during an actual event for definitive diagnosis .
   - **Convulsive Syncope**: Brief, irregular convulsive movements can occur

during syncope, along with incontinence. Usually shorter duration and less rhythmic than true seizures .

### Recommended Next Steps

**The gold standard for diagnosis is video-EEG monitoring** to capture a typical event . Other helpful investigations include:

- **Serum prolactin** drawn within 10-20 minutes of an event (elevated in GTCS, normal in PNES)
- **Cardiac evaluation** (ECG, Holter monitor) to rule out arrhythmic syncope
- Detailed history of **aura**, **postictal confusion**, and **injury patterns** (tongue biting strongly suggests GTCS)

**Conclusion:** While the normal investigations are reassuring, the clinical description is most consistent with generalized tonic-clonic seizures. The diagnosis is **provisional** pending capture of an event on video-EEG monitoring, which is essential for definitive diagnosis and appropriate treatment planning .

Based on the clinical presentation of a 32-year-old male with episodes of falling, uncontrollable shaking, loss of consciousness, and urinary incontinence—combined with normal EEG and MRI findings—here is the likely diagnostic approach:

### Primary Diagnostic Considerations

#### 1. **Psychogenic Non-Epileptic Seizures (PNES) - Most Likely**
The normal EEG and MRI findings, combined with the typical seizure-like presentation, strongly suggest PNES as the leading consideration. Key supporting evidence:

- **EEG specificity**: PNES patients show **no abnormal electrical activity** on EEG, which is the most useful test for distinguishing these from epileptic seizures
- **Symptom overlap**: PNES can present with convulsions, loss of consciousness, urinary incontinence, and tongue biting—mimicking epileptic seizures
- **Prevalence**: PNES is commonly associated with **mental stress, emotional pain, or physical conditions** rather than brain electrical abnormalities
- **Demographic fit**: While more common in women, PNES can affect men, particularly those in high-stress professions

#### 2. **Epilepsy (Tonic-Clonic Seizures) - Still Possible Despite Normal Tests**
The clinical description matches tonic-clonic seizures, but normal neuroimaging and EEG do not definitively rule out epilepsy:

- **Normal interictal EEG**: Approximately 50% of people with epilepsy may have normal EEGs between seizures
- **Urinary incontinence correlation**: Studies show ~39% of epilepsy patients experience urinary incontinence, most commonly with tonic-clonic seizures due to increased intra-abdominal pressure and pelvic floor muscle contractions
- **Diagnostic challenge**: The presentation could represent **early or occult epilepsy** where abnormalities haven't been captured

#### 3. **Syncope with Convulsive Activity - Less Likely**
Though possible, this is less probable given the full description:

- **Key distinguishing factor**: While syncope can cause urinary incontinence, convulsive syncope typically lacks the prolonged postictal confusion seen in seizures
- **Cardiac evaluation needed**: If syncope is suspected, cardiac monitoring (ECG, Holter) would be essential to rule out arrhythmias

### Critical Diagnostic Limitation of Urinary Incontinence

Importantly, **urinary incontinence has poor discriminatory value** between epileptic and non-epileptic events. A systematic review found:
- Sensitivity: 38%
- Specificity: 57%
- Positive likelihood ratio: 0.879 (no diagnostic value)
**Conclusion**: Incontinence cannot reliably distinguish seizure types and should not heavily influence the differential diagnosis

### Recommended Next Steps

The diagnosis requires **comprehensive evaluation** beyond initial EEG/MRI:

1. **Video-EEG monitoring**: Capture typical episodes to correlate clinical semiology with brain activity
2. **Psychiatric/psychological assessment**: Screen for stress, anxiety, depression, or trauma history
3. **Cardiac workup**: ECG and Holter monitoring to exclude arrhythmogenic syncope
4. **Detailed seizure history**: Document duration, triggers, prodromal symptoms, and postictal recovery
5. **Serum prolactin**: If drawn within 20 minutes of an event, can help differentiate epileptic from psychogenic seizures

### Final Assessment

Given the **normal EEG and MRI** in the setting of stereotyped seizure-like episodes, **Psychogenic Non-Epileptic Seizures (PNES)** is the most likely diagnosis. However, epilepsy cannot be definitively excluded without long-term monitoring. The occupational stress of banking and the patient's geographic location (Texas - suggesting possible differential access to specialized epilepsy centers) should also be considered in the comprehensive evaluation.

Based on the clinical presentation, the **most likely diagnosis is generalized tonic-clonic (GTC) seizures (epilepsy)**, though **psychogenic nonepileptic seizures (PNES)** must be strongly considered given the normal EEG and MRI.

#### Key Diagnostic Considerations

**Evidence Supporting Epileptic Seizures:**
- Classic semiology: sudden loss of consciousness, falling, convulsive shaking, and urinary incontinence are hallmark features of GTC seizures
- Urinary incontinence occurs in approximately 39% of patients with epilepsy, particularly during GTC seizures due to pelvic muscle contractions and increased abdominal pressure

**Evidence Against Epilepsy (Raising Suspicion for PNES):**
- **Normal EEG and MRI**: While these don't exclude epilepsy (up to 50% of patients have normal interictal EEGs), they shift probability toward non-epileptic events
- **Normal imaging** makes structural epilepsy less likely
- Urinary incontinence has poor discriminatory value, occurring in both epileptic and non-epileptic events (sensitivity 38%, specificity 57%)

#### Critical Differentiating Features to Assess

**If PNES:**
- Episodes often last **longer** (several minutes) with **waxing/waning intensity**
- **Minimal or no postictal confusion** (vs. >10 minutes in GTC seizures)
- **Atypical motor patterns**: side-to-side head movements, pelvic thrusting, asynchronous limb movements
- Patients may **actively resist eye opening** and maintain normal vital signs

**If Convulsive Syncope:**
- **Brief duration** (seconds) with **irregular, non-rhythmic jerks**
- **Prodromal symptoms**: nausea, lightheadedness, vision changes, pallor
- **Very brief confusion** (<10 minutes) post-event
- Often triggered by pain, emotion, or prolonged standing

#### Recommended Next Steps

1. **Video-EEG monitoring**: The gold standard to capture a typical event and definitively differentiate epileptic from non-epileptic seizures
2. **Detailed history**: Ask about episode duration, tongue biting (lateral vs. tip), postictal confusion, and triggers
3. **Cardiac evaluation**: Rule out convulsive syncope (ECG, Holter monitor) as this can present similarly

**Conclusion**: While GTC epilepsy remains the leading diagnosis due to the classic convulsive semiology, the normal investigations warrant **high suspicion for PNES** until proven otherwise with video-EEG capture of a typical event.

Based on the clinical presentation, the most likely diagnosis is **generalized tonic-clonic seizures (grand mal seizures)** as part of a probable epilepsy syndrome, despite the normal EEG and MRI findings.

### Diagnostic Rationale

The clinical features strongly suggest tonic-clonic seizures:

- **Falling and shaking uncontrollably** matches the tonic phase (sudden muscle stiffening causing falls) followed by the clonic phase (rhythmic jerking movements) characteristic of tonic-clonic seizures
- **Loss of consciousness** is a defining feature of tonic-clonic seizures, occurring during both the tonic and clonic phases
- **Urinary incontinence** commonly occurs at the end of generalized tonic-clonic seizures due to bladder muscle contraction and sphincter relaxation

### Significance of Normal EEG and MRI

A normal EEG does **not** exclude epilepsy:
- EEG recordings may be normal between seizures (interictal period) in people with epilepsy
- Scalp EEG can miss seizure activity, especially if originating from deep brain structures
- After a single seizure, "sometimes the diagnosis of epilepsy can't be made" even with testing

A normal MRI is also consistent with epilepsy:
- Many epilepsy cases have no identifiable structural abnormality on imaging
- MRI is primarily used to rule out other causes such as stroke, tumors, or structural lesions

### Key Differential Diagnoses to Exclude

**Atonic seizures** are less likely because they involve sudden loss of muscle tone without the convulsive shaking phase . While they cause falls, consciousness is typically preserved .

**Psychogenic non-epileptic seizures (PNES)** are unlikely given the presence of urinary incontinence and classic postictal confusion (though the latter wasn't specifically mentioned).

### Recommended Next Steps

The patient should be referred to a neurologist or epileptologist for:
1. **Video-EEG monitoring** to capture ictal activity during an event
2. **Cardiac evaluation** (ECG/Holter) to rule out convulsive syncope
3. **Laboratory testing** to exclude metabolic causes
4. **Detailed seizure journaling** documenting triggers, duration, and postictal symptoms

The diagnosis remains clinical despite normal initial tests, and treatment with anti-seizure medication may be warranted given the high risk of injury from these events .

The presentation of a 32-year-old male with episodes of falling, shaking, loss of consciousness, and urinary incontinence, but with normal EEG and MRI, raises several diagnostic possibilities. Let me search for more information about seizures with normal neuroimaging and EEG findings to provide a comprehensive differential diagnosis.
Based on the clinical presentation of a 32-year-old male with episodes of falling, shaking, loss of consciousness, and urinary incontinence, but with normal EEG and MRI, the most likely diagnosis is **psychogenic non-epileptic seizures (PNES)**, also known as functional seizures or dissociative seizures.

### Key Diagnostic Features Supporting PNES:

#### Clinical Presentation
- **Convulsive activity with preserved consciousness**: PNES can present with jerky, rhythmic, or twitching motions that resemble epileptic seizures, including falling down, loss of consciousness, and urinary incontinence
- **Normal EEG**: This is a critical feature - while routine EEG has limitations, patients with PNES do not show epileptiform abnormalities during their events, unlike true epileptic seizures

#### Epidemiology
- PNES accounts for **20-30% of patients referred to epilepsy centers** for refractory seizures
- Common in adults with high-stress occupations or significant life stressors

#### Why Not Other Diagnoses?

**Atonic seizures**: While these cause falls, they involve sudden loss of muscle tone (going limp) rather than "shaking uncontrollably". They also typically last <15 seconds and may show specific EEG patterns.

**Epileptic tonic-clonic seizures**: These would almost always show EEG abnormalities during or between seizures, especially with multiple episodes.

**Convulsive syncope**: Though possible, the stereotyped recurrence and lack of cardiovascular triggers make this less likely.

### Recommended Evaluation:

1. **Video-EEG monitoring**: Gold standard for diagnosis - capturing a typical event to confirm absence of epileptiform activity during the spell
2. **Psychiatric/psychological assessment**: To identify underlying stressors, trauma, or psychiatric comorbidities
3. **Laboratory studies**: Rule out metabolic/toxic causes
4. **Cardiac evaluation**: Consider Holter monitor to exclude arrhythmia

**Important distinction**: PNES are **involuntary** and not consciously feigned - they represent a complex physical reaction to psychological stress, not intentional malingering.

The normal neuroimaging and EEG, combined with convulsive semiology, strongly suggest PNES rather than epilepsy. However, definitive diagnosis requires video-EEG confirmation of a typical event.
Based on the clinical presentation and normal diagnostic studies, the **most likely diagnosis is Psychogenic Non-Epileptic Seizures (PNES)**.
**Key Clinical Features Supporting PNES:**

1. **Normal EEG and MRI:** This is the most critical finding. While interictal EEG can be normal in up to 50% of epilepsy patients, the combination of completely normal EEG and MRI makes epilepsy less likely, whereas these studies are typically normal in PNES.
2. Convulsive activity with loss of consciousness and incontinence: PNES can mimic tonic-clonic seizures, including shaking movements, falling, loss of consciousness, and urinary incontinence. These episodes are

often clinically indistinguishable from epileptic seizures without EEG monitoring.
3. Demographic context: While PNES is more common in women, it can occur in men. Stress-related factors (common in high-pressure banking careers) can be triggers.

**Important Considerations:**

**Epilepsy is NOT ruled out:**

- Approximately 30% of patients with temporal lobe epilepsy can have normal MRI
- Interictal EEGs may miss epileptiform activity
- "Urinary incontinence has no discriminatory value" between epileptic and non-epileptic events

Critical next steps:

1. Video-EEG monitoring: The gold standard for definitive diagnosis - captures the actual event to confirm absence of epileptiform activity during convulsive episodes
2. Cardiac evaluation: Must rule out syncope (particularly convulsive syncope) which can present similarly:
    - ECG and Holter monitoring
    - One case report specifically noted "urinary incontinence without convulsion" may indicate cardiogenic syncope rather than epilepsy
3. Psychological assessment: To identify underlying stressors, trauma, or mental health conditions often associated with PNES

**Bottom Line:**

The normal EEG and MRI in the setting of convulsive episodes strongly suggest **PNES** as the working diagnosis, but this must be confirmed with video-EEG monitoring while ruling out cardiac causes of syncope before finalizing the diagnosis.
