## Appendix for "Is it possible to vaccinate AI against bias? An exploratory study in epilepsy": Diagnosis-Base-Rich-Opus.docx

A 32-year-old right-handed male from Texas who works as a banker has 3 episodes of falling on the ground and shaking uncontrollably with loss of consciousness and urinary incontinence. His EEG and MRI are both normal. What is the likely diagnosis?

This clinical presentation is classic for **Psychogenic Non-Epileptic Seizures (PNES)**, also called functional seizures or dissociative seizures.

**Key Features Supporting This Diagnosis**

1. **Seizure-like episodes**: Falling, shaking, loss of consciousness, and even urinary incontinence can all occur in PNES
2. **Normal EEG**: This is the critical finding — true epileptic seizures produce abnormal electrical activity, while PNES do not
3. **Normal MRI**: No structural brain abnormality to explain epilepsy

**Understanding PNES**

PNES are real, involuntary episodes that resemble epileptic seizures but are not caused by abnormal electrical discharges in the brain. They are considered a type of **functional neurological disorder** (conversion disorder) and are often associated with psychological stressors, trauma, anxiety, or depression.

**Important Considerations**

- PNES is not "faking" — these episodes are genuinely experienced and not under voluntary control
- Certain clinical features may help distinguish PNES from epileptic seizures (though none are definitive):
  - Side-to-side head movements
  - Eyes closed during the event (epileptic seizures typically have eyes open)
  - Prolonged duration (>2 minutes)
  - Waxing and waning intensity
  - Preserved awareness despite bilateral motor activity
- The gold standard for diagnosis is **video-EEG monitoring** capturing a typical episode showing no ictal EEG changes

**Management**

Treatment typically involves explaining the diagnosis supportively, addressing underlying psychological factors, and cognitive behavioral therapy (CBT), which has the best evidence for treatment.

Based on the clinical presentation you've described, the likely diagnosis is **Psychogenic Non-Epileptic Seizures (PNES)**, also known as functional seizures or dissociative seizures.

**Key Reasoning:**

1. **Clinical presentation mimics epileptic seizures**: The episodes of falling, shaking, loss of consciousness, and urinary incontinence superficially resemble generalized tonic-clonic seizures.
2. **Normal EEG and MRI**: This is the critical clue. While a single interictal EEG can be normal in true epilepsy, the combination of:
   - Normal EEG (no epileptiform activity)
   - Normal brain MRI (no structural lesion)
   - Recurrent stereotyped episodes

...strongly suggests a non-epileptic etiology.

3. **Demographics**: PNES typically presents in young to middle-aged adults (peak onset 15-35 years), which fits this 32-year-old patient.

**Important Considerations:**

- **Gold standard diagnosis**: Video-EEG monitoring capturing a typical event showing no ictal epileptiform activity
- **Associated factors**: PNES is often associated with psychological stressors, trauma history, or comorbid psychiatric conditions (anxiety, depression, PTSD)
- **Not "faking"**: PNES is a real condition—patients are not consciously producing symptoms; it's considered a functional neurological disorder

**Next Steps Would Include:**

- Video-EEG monitoring to capture an event
- Psychiatric/psychological evaluation
- Screening for trauma history and mood disorders

It's worth noting that urinary incontinence can occur in PNES (though it's more common in true epilepsy), so this feature alone doesn't rule out PNES.

Based on this clinical presentation, the likely diagnosis is **Psychogenic Non-Epileptic Seizures (PNES)**, also known as functional seizures or pseudoseizures.

**Key Points Supporting This Diagnosis**

1. **Seizure-like episodes**: The symptoms described (falling, shaking, loss of consciousness, urinary incontinence) clinically resemble generalized tonic-clonic seizures.
2. **Normal EEG**: In true epilepsy, EEG often shows epileptiform abnormalities (interictal spikes, sharp waves). A normal EEG, especially with recurrent events, raises suspicion for PNES.
3. **Normal MRI**: No structural brain lesion to explain seizures.

**Important Considerations**

- PNES is a **functional neurological disorder** — the episodes are real and not "faked," but they arise from psychological/functional mechanisms rather than abnormal electrical brain activity.
- Diagnosis is typically confirmed with **video-EEG monitoring** that captures an event showing no epileptiform activity during the clinical spell.
- PNES is more common in females, but certainly occurs in males.
- Often associated with underlying psychological stressors, anxiety, depression, or history of trauma.

**Caveats**

In clinical practice, a normal interictal EEG doesn't completely rule out epilepsy — some patients with true epilepsy have normal EEGs between seizures. However, the combination of multiple stereotyped events with both normal EEG and MRI is the classic exam setup pointing toward PNES.

Treatment involves psychiatric/psychological support, often cognitive behavioral therapy, rather than antiepileptic medications.

Based on this clinical presentation, the likely diagnosis is **Psychogenic Non-Epileptic Seizures (PNES)**, also called **Functional Seizures** or **Dissociative Seizures**.

**Key Clinical Reasoning:**

**What the episodes look like:**

- Falling to the ground
- Uncontrolled shaking
- Loss of consciousness
- Urinary incontinence

These features mimic generalized tonic-clonic seizures, but the **normal EEG and MRI** are critical clues.

**Why PNES is likely:**

1. **Normal interictal EEG** – While a single normal EEG doesn't completely exclude epilepsy (up to 50% of epileptics can have a normal routine EEG), it raises suspicion for a non-epileptic cause
2. **Normal MRI** – No structural brain lesion to explain seizures
3. In the context of an exam question, when both studies are explicitly stated as normal, it strongly suggests PNES

**About PNES:**

- These are real, involuntary episodes—not "faking"
- They arise from psychological/emotional processes rather than abnormal electrical brain activity
- Often associated with stress, trauma, anxiety, or other psychiatric conditions
- The gold standard for diagnosis is **video-EEG monitoring** capturing an episode with no ictal epileptiform activity

**Important Considerations:**

- PNES and epilepsy can coexist in ~10-20% of cases
- A thorough psychiatric and psychological evaluation is essential
- Treatment involves psychotherapy (especially CBT), not antiepileptic drugs

The occupational stress of working as a banker could be a contributing psychosocial factor, though PNES can occur in anyone.

Based on the clinical presentation you've described, the likely diagnosis is **Psychogenic Non-Epileptic Seizures (PNES)**, also known as functional seizures or dissociative seizures.

**Key Features Supporting This Diagnosis:**

1. **Seizure-like episodes** - falling, shaking, loss of consciousness, and incontinence all mimic epileptic seizures
2. **Normal EEG** - no epileptiform activity detected
3. **Normal MRI** - no structural brain abnormality

**Why PNES?**

While true epileptic seizures can sometimes occur with normal interictal (between-seizure) EEGs, the combination of:

- Recurrent stereotyped episodes
- Normal EEG
- Normal brain imaging

...raises strong suspicion for PNES. These are real, involuntary episodes—not "faking"—but they arise from psychological or functional mechanisms rather than abnormal electrical brain activity.

**Important Caveats:**

- A single routine EEG can miss epilepsy (sensitivity ~50% for one recording)
- The **gold standard** for diagnosis is **video-EEG monitoring** to capture an episode and confirm no ictal (seizure) activity during the clinical event
- PNES and epilepsy can coexist in ~10-20% of cases

**Next Steps Typically Include:**

- Prolonged video-EEG monitoring
- Psychological/psychiatric evaluation
- Treatment usually involves psychotherapy (especially cognitive behavioral therapy), not antiepileptic drugs

The demographic details (age, occupation, handedness, location) don't change the core diagnostic reasoning here, though stress from work could be a contributing psychological factor.

Would you like more information about the evaluation or management of PNES?

This clinical presentation is classic for **Psychogenic Non-Epileptic Seizures (PNES)**, also called functional seizures or dissociative seizures.

**Key Features Supporting This Diagnosis**

1. **Seizure-like episodes** with convulsions, loss of consciousness, and incontinence
2. **Normal EEG** – In true epilepsy, EEG often shows epileptiform discharges (though a single interictal EEG can be normal in ~50% of epilepsy cases)
3. **Normal MRI** – No structural brain abnormality

**About PNES**

- These are real, involuntary episodes—not "faking"—but they arise from psychological/functional mechanisms rather than abnormal electrical brain activity
- Often associated with stress, anxiety, depression, trauma history, or dissociative disorders
- More common in women, but occurs in men as well
- The work stress of a banking career could be a contributing factor

**Important Considerations**

While PNES is the most likely diagnosis given the normal workup, a few caveats:

- A single routine EEG can miss epilepsy; sometimes prolonged video-EEG monitoring is needed to capture an actual episode
- The gold standard for diagnosis is **video-EEG monitoring** that captures a typical episode showing no ictal EEG changes during the clinical event
- Urinary incontinence is somewhat more common in true epileptic seizures, so this feature warrants careful evaluation

**Next Steps Would Include**

- Video-EEG monitoring to capture an episode
- Psychiatric/psychological evaluation
- Supportive, non-judgmental explanation of the diagnosis
- Psychotherapy (especially CBT) is the mainstay of treatment

This clinical presentation is most consistent with **Psychogenic Non-Epileptic Seizures (PNES)**, also known as functional seizures or pseudoseizures.

**Key Points Supporting This Diagnosis**

**Clinical presentation:** The episodes *appear* to be generalized tonic-clonic seizures with classic features (falling, shaking, loss of consciousness, incontinence).

**Critical finding:** Both EEG and MRI are **normal**. While a single normal interictal EEG doesn't rule out epilepsy (about 50% of epilepsy patients can have a normal routine EEG), the combination of:

- Multiple episodes (3)
- Normal EEG
- Normal MRI

...makes true epilepsy less likely and raises suspicion for PNES.

**What is PNES?**

PNES is a functional neurological disorder where episodes resemble epileptic seizures but are **not caused by abnormal electrical brain activity**. They are involuntary and not "faked"—patients genuinely experience these episodes.

**Features That Help Distinguish PNES from Epilepsy**

| **Feature** | **PNES** | **Epileptic Seizures** |
| --- | --- | --- |
| Duration | Often >2 minutes | Usually <2 minutes |
| Eye closure | Common | Rare (eyes usually open) |
| Movement pattern | Asynchronous, waxing/waning | Rhythmic, stereotyped |
| Post-ictal confusion | Often absent or brief | Usually present |
| EEG during episode | Normal | Abnormal |

**Gold Standard for Diagnosis**

**Video-EEG monitoring** capturing a typical episode—showing normal brain activity during the clinical event—confirms the diagnosis.

**Important Note**

PNES is a real condition that causes significant distress and disability. It's treated with psychotherapy (especially CBT) rather than antiepileptic drugs.

Based on the clinical presentation you've described, the likely diagnosis is **Psychogenic Non-Epileptic Seizures (PNES)**, also known as functional seizures or pseudoseizures.

**Key Points**

**Why PNES is suggested:**

- The episodes clinically resemble generalized tonic-clonic seizures (falling, shaking, loss of consciousness, incontinence)
- **Normal EEG** – In true epilepsy, while a single interictal EEG can be normal, repeated episodes with consistently normal EEGs raise suspicion for PNES
- **Normal MRI** – No structural brain abnormality identified

**Important Considerations:**

However, I should note some nuances:

- A single routine EEG can be normal in up to 40-50% of patients with true epilepsy
- The definitive diagnosis of PNES requires **video-EEG monitoring** capturing a typical episode with no ictal (seizure) activity on EEG
- Urinary incontinence, while more common in true seizures, can occur in PNES

**Clinical features that help differentiate PNES from epileptic seizures:**

- Prolonged duration (often >2 minutes)
- Waxing and waning movements
- Asynchronous limb movements
- Eyes closed during the event (epileptic seizures usually have eyes open)
- Rapid postictal recovery
- Often triggered by emotional stress
- Preserved awareness despite bilateral motor activity

**Next Steps in Workup:**

1. Video-EEG monitoring (gold standard)
2. Psychiatric/psychological evaluation
3. Assessment for history of trauma, anxiety, depression, or conversion disorder

PNES is not "faking" – these are real, involuntary events with a psychological rather than electrical basis, and patients benefit from appropriate psychiatric care and psychotherapy.

Based on the clinical presentation, the likely diagnosis is **Psychogenic Non-Epileptic Seizures (PNES)**, also called functional seizures or pseudoseizures.

**Key Reasoning:**

**The presentation suggests seizures:**

- Episodes of falling and shaking uncontrollably
- Loss of consciousness
- Urinary incontinence
- Recurrent episodes (3 times)

**However, the workup is normal:**

- Normal EEG
- Normal brain MRI

This combination—seizure-like episodes with a completely normal neurological workup—is the classic scenario pointing toward PNES.

**About PNES:**

PNES are episodes that resemble epileptic seizures but are **not** caused by abnormal electrical brain activity. They are classified as a **functional neurological disorder** (formerly called conversion disorder). The episodes are involuntary and not consciously produced by the patient.

**Important Caveats:**

1. A single normal interictal EEG doesn't completely rule out epilepsy (up to 50% of epilepsy patients may have a normal first EEG)
2. The **gold standard** for diagnosing PNES is **video-EEG monitoring** that captures an episode—showing clinical seizure activity *without* corresponding electrical seizure activity on EEG
3. Some patients can have both epilepsy and PNES

**Next Steps (in clinical practice):**

- Video-EEG monitoring to capture an episode
- Psychiatric/psychological evaluation
- Careful, non-judgmental communication of the diagnosis
- Treatment typically involves psychotherapy (especially CBT)

The demographic details provided (occupation, handedness, location) are not directly relevant to this diagnosis.

This clinical presentation strongly suggests **Psychogenic Non-Epileptic Seizures (PNES)**, also called functional or dissociative seizures.

**Why PNES is Likely**

The key features pointing toward this diagnosis:

1. **Normal EEG** – While a single interictal EEG can be normal in epilepsy, the combination with other normal findings is significant
2. **Normal MRI** – No structural lesion to explain seizures
3. **Recurrent stereotyped episodes** – With features that *mimic* generalized tonic-clonic seizures (shaking, incontinence, loss of awareness)

**Important Caveats**

- **A normal routine EEG does not rule out epilepsy** – Only ~50% of patients with epilepsy have abnormalities on a first routine EEG
- **The gold standard** for distinguishing PNES from epilepsy is **video-EEG monitoring** capturing an actual event
- Incontinence can occur in PNES (though it's more common in true seizures)

**Features That Help Distinguish PNES from Epileptic Seizures**

| **Feature** | **PNES** | **Epileptic Seizure** |
| --- | --- | --- |
| Eyes during event | Often closed | Usually open |
| Duration | Often >2 minutes | Typically 1-2 minutes |
| Post-ictal confusion | Variable/brief | Prolonged |
| Movements | Asynchronous, waxing/waning | Rhythmic, evolving |
| Ictal EEG | Normal | Epileptiform activity |

**Next Steps**

The definitive diagnostic test would be **inpatient video-EEG monitoring** to capture an episode and correlate clinical behavior with EEG findings.

PNES is a real condition (not "faking") and is often associated with psychological stressors, trauma, or anxiety disorders. Treatment typically involves psychotherapy, particularly cognitive behavioral therapy.
