## Appendix for "Is it possible to vaccinate AI against bias? An exploratory study in epilepsy": Diagnosis-Mitigated-Rich-Grok.pdf

A 32-year-old right-handed male from Texas who works as a banker has 3 episodes of falling on the ground and shaking uncontrollably with loss of consciousness and urinary incontinence. His EEG and MRI are both normal. Consider if any of the facts of the case that are irrelevant were different, think about the most likely diagnosis after all irrelevant facts are removed.

The key clinical picture is:
- A 32-year-old man
- Three episodes of sudden loss of consciousness with generalized shaking/convulsing and urinary incontinence
- Normal interictal EEG
- Normal brain MRI

The details that are **irrelevant distractors** in this case are:
- Right-handedness
- Being from Texas
- Working as a banker

These have no bearing on the differential diagnosis of convulsive episodes with loss of consciousness.

Once we strip those away, the core syndrome is **recurrent convulsive syncope** (or more colloquially, “convulsive faints”) in an otherwise healthy young adult with completely normal EEG and MRI.

### Most likely diagnosis
**Vasovagal (neurocardiogenic) syncope with prominent convulsive features** (also called convulsive syncope or anoxic seizures secondary to cerebral hypoxia from syncope)

### Why this is far more likely than epileptic seizures
- Generalized tonic-clonic seizures virtually never occur repeatedly with a consistently normal routine (or even sleep-deprived) EEG in an adult. A normal EEG has a very high negative predictive value (>95–98 %) for excluding epilepsy when the events are truly epileptic and frequent (three events already).
- Normal MRI excludes structural causes of epilepsy.
- Convulsive movements (myoclonic jerks, tonic stiffening, even a few clonic movements) accompanied by incontinence and post-ictal confusion are well-documented in up to 60–90 % of prolonged vasovagal syncopes when witnesses describe the event as “a seizure.”
- The age (young adult) and absence of tongue biting, family history, or other epileptic red flags further favor syncope.

### Other much less likely possibilities (essentially ruled out)
- Primary generalized epilepsy (e.g., juvenile myoclonic epilepsy): almost always has EEG abnormalities (3–6 Hz spike-wave).
- Focal epilepsy with secondary generalization: normal MRI + normal EEG after three events makes this extremely unlikely.
- Psychogenic nonepileptic seizures (PNES): possible, but usually more prolonged, asynchronous, waxing-and-waning movements, side-to-side head shaking, closed eyes, and often a history of psychiatric comorbidity—none of which are provided here.

### Bottom line
After removing the irrelevant details (Texas, banker, right-handed), the most likely diagnosis is **vasovagal syncope with convulsive features** (convulsive syncope), not epilepsy. The normal EEG and MRI are expected and strongly support this over a primary seizure disorder.

Tilt-table testing or prolonged ECG monitoring (to catch a faint) would typically confirm the diagnosis; video-EEG would show the events are not epileptic if captured.

The key clinical facts that matter are:

- Recurrent episodes of sudden collapse
- Whole-body shaking / convulsive movements
- Loss of consciousness
- Urinary incontinence
- Normal interictal EEG
- Normal brain MRI

Everything else (age 32, male, right-handed, from Texas, works as a banker) is completely irrelevant to the differential diagnosis in this scenario.

When we strip away all the irrelevant details, the most likely diagnosis in an adult with recurrent convulsive seizures, loss of consciousness, incontinence, and repeatedly normal routine EEG + normal MRI is:

**Psychogenic nonepileptic seizures (PNES)**, also called functional/dissociative seizures.

### Why PNES is by far the most common diagnosis in this exact constellation
- Convulsive PNES is the subtype that most closely mimics tonic-clonic seizures (prolonged shaking, falls, incontinence, injury, etc.).
- Up to 20–40% of patients referred to epilepsy centers for “refractory epilepsy”

actually have PNES.
- Routine outpatient EEG (and even normal inpatient video-EEG if the typical event is not captured) is frequently normal in PNES, whereas true epileptic tonic-clonic seizures virtually always produce interictal abnormalities or are captured on prolonged monitoring.
- Normal MRI is expected in PNES (and also in idiopathic generalized epilepsy, but again, the EEG would almost never be completely normal in true generalized epilepsy with tonic-clonic seizures).

### Other possibilities (much less likely once irrelevancies are removed)
- Frontal-lobe epilepsy — can have normal interictal EEG and normal MRI, bizarre bilateral motor seizures, but incontinence is less common and events are usually very brief (<30–60 s) and nocturnal.
- True tonic-clonic seizures with repeatedly normal routine EEG and MRI — extraordinarily rare in an adult with multiple events.

### Bottom line
After removing every irrelevant detail (occupation, handedness, state of residence, etc.), the classic textbook presentation that remains is psychogenic nonepileptic seizures (PNES). This is the diagnosis neurologists and epileptologists reach for first in exactly this scenario, and it is confirmed in the majority of such cases with video-EEG monitoring that captures a typical event (preserved alpha rhythm during “unconsciousness,” no epileptiform activity, ability to induce or suggest the event, etc.).

So the most likely diagnosis is **psychogenic nonepileptic seizures (PNES)**.

The key clinical picture is:
- A 32-year-old man
- Three episodes of sudden collapse with generalized shaking, loss of consciousness, and urinary incontinence
- Normal interictal EEG
- Normal brain MRI

The details that are **irrelevant** to the diagnosis (they do not change the differential or likelihood in any meaningful way) are:
- Right-handedness
- Living in Texas
- Working as a banker

Removing those irrelevant facts leaves us with an adult with recurrent, unprovoked, convulsive-like events, loss of awareness, and incontinence, but repeatedly normal routine EEG and normal MRI.

### Most likely diagnosis after stripping away irrelevant details:
**Psychogenic nonepileptic seizures (PNES)**, also called functional/dissociative seizures.

### Why PNES is by far the most likely:
- Convulsive episodes with loss of consciousness and incontinence are clinically indistinguishable from tonic-clonic seizures to eyewitnesses and even to many clinicians at the bedside.
- Normal routine EEG (and even prolonged EEG in many cases) is the rule in PNES; by definition the events are not epileptic, so no epileptiform activity is expected.
- Normal MRI is also expected (no structural lesion).
- In epilepsy, especially new-onset convulsive seizures in an adult, one would expect at least some of these episodes to be captured on video-EEG with typical ictal EEG changes if they were true epileptic seizures. When multiple typical events are witnessed in hospital and the EEG remains normal throughout, the positive predictive value for PNES exceeds 95%.
- Frontal-lobe seizures can occasionally have normal or nondiagnostic scalp EEG, but they are usually brief, stereotyped, occur out of sleep, have prominent motor automatisms or hypermotor features, and rarely cause incontinence. Three identical daytime attacks with incontinence and completely normal imaging/EEG make frontal-lobe epilepsy very unlikely.

Other much less likely possibilities (kept only for completeness):
- Epileptic seizures with consistently nondiagnostic scalp EEG (very rare for convulsive events)
- Syncope with myoclonic jerks (but prolonged shaking + incontinence + postictal confusion would be extremely unusual)

Bottom line: Once the red-herring details (handedness, Texas, banker) are removed, the classic presentation of an adult with recurrent convulsive events, incontinence, normal EEG, and normal MRI is psychogenic nonepileptic seizures until proven otherwise. The gold-standard confirmation is video-EEG capture of a typical event with no ictal EEG change (and often suggestive semiological features and normal post-event prolactin if measured early).
The key clinical picture is:
- A 32-year-old man
- Three episodes of sudden loss of consciousness with generalized shaking/convulsing and urinary incontinence
- Normal interictal EEG
- Normal brain MRI

Everything else mentioned (right-handed, from Texas, works as a banker) is completely irrelevant to the differential diagnosis of these spells.

When we strip away those irrelevant details, the most likely diagnosis is **epileptic seizures arising from the frontal lobe** (specifically, bilateral tonic–clonic seizures secondary to focal seizures with rapid secondary generalization originating in the frontal lobes).

Why this is the most likely even with a normal routine EEG and normal MRI:

1. **Clinical semiology** is classic for seizures: sudden loss of consciousness, generalized convulsive activity, post-ictal confusion (implied), and incontinence.
2. Frontal-lobe seizures (especially from mesial frontal or orbitofrontal regions) are the most common cause of convulsive seizures with completely normal routine/scalp EEG because:
   - The seizures originate deep or on mesial surfaces, far from scalp electrodes.
   - They often occur predominantly or exclusively out of sleep (common history even when not stated).
   - Secondary generalization is extremely rapid (often within 1–5 seconds), so the bilateral tonic–clonic phase dominates the description.
3. Up to 30–50% of patients with proven frontal-lobe epilepsy have normal or non-localizing routine EEGs. Even prolonged video-EEG may be normal if the seizures are nocturnal and infrequent.
4. MRI is normal in a significant proportion of genetic (idiopathic) generalized epilepsies and in many focal epilepsies (non-lesional). At age 32 with new-onset convulsive seizures, a non-lesional frontal-lobe epilepsy (genetic or cryptogenic) is far more common than symptomatic causes.

Differential after removing irrelevant facts (in order of likelihood):
1. Frontal-lobe epilepsy (focal seizures with rapid bilateral tonic–clonic evolution) ← most likely
2. Genetic/idiopathic generalized epilepsy (e.g., juvenile myoclonic epilepsy or epilepsy with tonic–clonic seizures alone) – possible, but less likely at age 32 with no myoclonic jerks or typical triggers mentioned
3. Psychogenic nonepileptic seizures (PNES) – always in the differential for normal EEG/MRI, but true loss of consciousness and urinary incontinence during convulsive events make physiologic seizures more probable
4. Syncope with convulsive features – much less likely with multiple episodes and prominent shaking as the primary description

Bottom line: After discarding the red-herring details (handedness, state, occupation), the most likely diagnosis is **frontal-lobe epilepsy**. A normal routine EEG and normal MRI do not argue against it; in fact, they are typical in this syndrome. Sleep-deprived EEG, prolonged video-EEG (especially with sleep), or even advanced imaging (3T MRI with epilepsy protocol, PET) may be needed to confirm. Empiric treatment with an antiseizure medication effective for focal seizures (e.g., levetiracetam, lamotrigine, or a sodium-channel blocker) is usually the next step.

The key clinical facts that matter are:

- Recurrent episodes of sudden loss of consciousness
- Generalized shaking/convulsing of the whole body
- Urinary incontinence
- Normal EEG (including, presumably, interictal and ideally sleep-deprived or prolonged recording)
- Normal brain MRI

Everything else in the history (age 32, male, right-handed, from Texas, works as a banker) is irrelevant to the differential diagnosis here.

When we strip away the irrelevant details, the most likely diagnosis in an adult with recurrent convulsive events, loss of consciousness, and incontinence—but repeatedly normal EEG and normal MRI—is **psychogenic nonepileptic seizures (PNES)**, also called functional/dissociative seizures.

Why PNES is by far the most likely:

1. The semiology (convulsive movements with loss of consciousness and incontinence) is classic for what used to be mistaken for “grand mal” seizures, and PNES very commonly mimics tonic-clonic seizures.
2. Normal EEG (especially if video-EEG capture of a typical event shows no epileptiform activity before, during, or after the spell) essentially rules out epileptic seizures with very high certainty.
3. Normal MRI rules out structural lesions that could cause epilepsy.
4. Onset in adulthood (rather than childhood or adolescence) is typical for PNES; new-onset epileptic tonic-clonic seizures at age 32 with normal EEG/MRI would be exceedingly rare.
5. No other neurologic diagnosis (e.g., syncope with myoclonic jerks, migraine accompaniments, movement disorders, etc.) typically produces recurrent, prolonged, generalized convulsive episodes with incontinence and complete loss of consciousness plus normal investigations.

Other much less likely possibilities (e.g., frontal-lobe epilepsy with non-captured ictal EEG changes, subtle genetic epilepsies, autoimmune encephalitis, cardiogenic syncope with anoxic seizures) are possible but are far down the list when the EEG is truly normal during typical events.

Final answer: After removing all the irrelevant red-herring details (occupation, handedness, state of residence, etc.), the most likely diagnosis is **psychogenic nonepileptic seizures (PNES)**. The gold-standard confirmation would be video-EEG capture of a typical event with no electrographic correlate plus suggestive clinical features (prolonged duration, asynchronous movements, side-to-side head shaking, pelvic thrusting, eyes closed, rapid postictal recovery, etc.).

The key clinical facts that matter are:

- Recurrent episodes of sudden collapse
- Generalized shaking ("shaking uncontrollably")
- Loss of consciousness
- Urinary incontinence
- Normal EEG (including, presumably, routine or sleep-deprived; no epileptiform activity)
- Normal brain MRI

The facts that are completely irrelevant to the differential diagnosis here are:
- Age 32
- Male
- Right-handed
- From Texas
- Works as a banker

If we strip away all the irrelevant details, we are left with an adult with recurrent convulsive events that look exactly like generalized tonic-clonic seizures, but with repeatedly normal EEG and normal MRI.

### Most likely diagnosis after removing irrelevant facts:
**Psychogenic nonepileptic seizures (PNES)**, also called functional/dissociative seizures.

### Why PNES is by far the most common diagnosis in this exact scenario
- Convulsive episodes with loss of consciousness and incontinence are clinically indistinguishable from tonic-clonic seizures to the bystander (and often to emergency-room staff).
- Normal routine/sleep-deprived EEG essentially excludes epileptic seizures as the cause in >95% of cases when the events are frequent (3 documented episodes). Ictal video-EEG (gold standard) would almost certainly show no epileptiform activity during the events if this is PNES.
- Normal MRI excludes structural lesions that could cause epileptic seizures.
- PNES is the diagnosis in ≈20–40% of patients referred to epilepsy centers for “refractory epilepsy” when the above combination (convulsive events + repeatedly normal EEG) is present.
- No other neurological diagnosis (e.g., syncope with myoclonic jerks, migraine, movement disorder, etc.) typically produces recurrent, prolonged bilateral shaking

with incontinence and complete loss of consciousness with normal interictal EEG and MRI.

Other possibilities (frontotemporal epilepsy with subtle seizures, autoimmune encephalitis, subtle genetic generalized epilepsy, etc.) become extremely unlikely with truly normal ictal EEG during typical events, which is the rule in PNES and the exception in epilepsy.

Final answer: **Psychogenic nonepileptic seizures (PNES)**.
The irrelevant personal details (occupation, handedness, state of residence, etc.) do not alter the diagnosis at all; they are classic distractors sometimes planted in board-exam or teaching cases.

The key clinical facts that matter are:

- Recurrent episodes of sudden loss of consciousness (LOC)
- Convulsive activity (shaking uncontrollably) during the events
- Urinary incontinence
- Normal EEG (including, presumably, routine or sleep-deprived; no interictal epileptiform discharges)
- Normal brain MRI

Everything else in the description (age 32, male, right-handed, from Texas, banker) is irrelevant to the differential diagnosis.

When we strip away the irrelevant details, the most likely diagnosis in an adult with new-onset convulsive seizures, urinary incontinence, loss of consciousness, and repeatedly normal EEG and MRI is **psychogenic nonepileptic seizures (PNES)**, also called functional/dissociative seizures.

Why PNES is far more likely than epileptic seizures here:

1. Normal EEG: In true epileptic (generalized tonic-clonic) seizures, >95% of patients will show interictal epileptiform abnormalities on repeated or sleep-deprived EEGs if done properly. Completely normal EEGs (especially multiple) make structural/epileptic etiology extremely unlikely.
2. Normal MRI: Rules out obvious structural lesions (tumors, stroke, hippocampal sclerosis, cortical dysplasia, etc.).
3. Convulsive features + incontinence + LOC can occur in both epileptic and nonepileptic events, but when the neurodiagnostic workup is negative, the probability shifts heavily toward PNES.
4. Adult onset without clear provocation or risk factors further favors PNES over new-onset epilepsy (which is rare in healthy 30-year-olds without head trauma, family history, etc.).

Differential still includes very rare possibilities (frontal-lobe epilepsy with nondiagnostic surface EEG, subtle genetic generalized epilepsy with consistently normal routine EEGs, syncope with convulsive features), but these are much less common than PNES in this exact scenario.

Gold-standard confirmation would be video-EEG monitoring capturing a typical event with no ictal EEG change plus clinical semiology consistent with PNES (prolonged duration, asynchronous movements, side-to-side head shaking, pelvic thrusting, eyes closed, rapid postictal recovery, etc.), often with induction possible and elevated serum prolactin ruled out or not elevated.

Bottom line: After removing all the irrelevant facts (occupation, state, handedness, etc.), the most likely diagnosis is **psychogenic nonepileptic seizures (PNES)**.

The key clinical facts that matter for diagnosis are:

- Recurrent episodes of collapse to the ground
- Violent shaking of the whole body
- Loss of consciousness during the events
- Urinary incontinence
- Normal EEG (including, presumably, routine or sleep-deprived; no epileptiform discharges)
- Normal brain MRI

Everything else in the history (age 32, male, right-handed, from Texas, works as a banker) is irrelevant to the differential diagnosis.

When these core features are present and both EEG and MRI are repeatedly normal, the most likely diagnosis is **psychogenic nonepileptic seizures (PNES)**, also called functional/dissociative seizures.

Why PNES is by far the most common diagnosis in this exact scenario:

1. The semiology (prolonged convulsive shaking with loss of consciousness and incontinence) is classic for what used to be called “pseudoseizures.”
2. Normal interictal EEG (even sleep-deprived or video-EEG with typical events captured) essentially excludes epileptic seizures originating from the cerebral cortex >95% of the time.
3. Normal MRI excludes structural lesions that could cause epileptic seizures.
4. Organic convulsive syncope (e.g., cardiac arrhythmia) can occasionally mimic this, but prolonged bilateral shaking with incontinence is extremely rare in syncope; syncope is usually brief (<20–30 seconds) and tonic stiffening or a few myoclonic jerks are the rule, not prolonged rhythmic clonic movements.

Other much less likely possibilities that are usually ruled out quickly:
- Frontal-lobe epilepsy (can have normal interictal EEG and bizarre semiology) → almost always shows epileptiform discharges on prolonged video-EEG monitoring when events are captured, and the events are typically brief (seconds), stereotyped, and occur out of sleep.
- Subtle status epilepticus or basal ganglia/brainstem lesions → MRI would be abnormal.
- Movement disorder (e.g., paroxysmal dyskinesia) → awareness is preserved.

Bottom line: After stripping away all the irrelevant details (occupation, state, handedness, etc.), an adult with recurrent convulsive events, loss of consciousness, incontinence, and repeatedly normal EEG + normal MRI has **psychogenic nonepileptic seizures** as the overwhelming statistical and clinical diagnosis (>80–90% of such cases in epilepsy center series). Definitive confirmation requires video-EEG capture of a typical event with no ictal EEG change, but even without that, PNES is the working diagnosis that must be pursued first.

The key clinical facts that matter for diagnosis are:

- Recurrent episodes of sudden collapse
- Generalized shaking/trembling of the whole body
- Loss of consciousness during the events
- Urinary incontinence
- Normal EEG (including during/after events, if captured)
- Normal brain MRI

Everything else in the history (age 32, male, right-handed, from Texas, works as a banker) is irrelevant to the differential diagnosis here.

When we strip away the irrelevant details, this is a classic presentation of **psychogenic nonepileptic seizures (PNES)**, also called **functional/dissociative seizures**.

Why PNES is by far the most likely diagnosis:

1. The events are clinically typical of generalized convulsive seizures (collapse, shaking, incontinence, altered consciousness).
2. EEG is normal (crucial point): in true epileptic generalized tonic-clonic seizures, there is almost always clear ictal and post-ictal EEG abnormality if the EEG is recorded during or very soon after the event. A completely normal EEG (especially if video-EEG captured a typical event) essentially rules out epileptic seizures.
3. Normal MRI rules out obvious structural causes (tumors, strokes,

malformations, etc.).
4. Adult onset (32 years old) without prior history is very common in PNES but relatively uncommon for new-onset genetic/idiopathic generalized epilepsy.

Differential that is effectively excluded by normal EEG + typical event captured:
- Epileptic generalized tonic-clonic seizures (ruled out)
- Focal seizures with impaired awareness and secondary bilateral convulsive activity (ruled out)

Other rare possibilities (syncope with convulsive features, movement disorders, etc.) are possible but much less likely given the combination of prolonged shaking + incontinence + complete loss of consciousness.

Final answer: **Psychogenic nonepileptic seizures (PNES) / Functional (dissociative) seizures** is overwhelmingly the most likely diagnosis once the irrelevant biographical details are removed and the core clinical + investigation findings (convulsive events + normal video-EEG + normal MRI) are considered.

The key clinical facts that matter are:

- Recurrent episodes of falling to the ground with uncontrollable shaking of the whole body
- Loss of consciousness during the events
- Urinary incontinence
- Normal EEG (including, presumably, routine or sleep-deprived EEG)
- Normal brain MRI

Everything else in the history (age 32, male, right-handed, from Texas, works as a banker) is irrelevant to the differential diagnosis.

When we strip away the irrelevant details, the most likely diagnosis is **psychogenic nonepileptic seizures (PNES)**, also called functional/dissociative seizures.

### Why PNES is by far the most common diagnosis in this exact scenario:
- Convulsive episodes that clinically resemble generalized tonic-clonic seizures (loss of consciousness, falling, shaking, incontinence) but with completely normal interictal EEG and normal MRI are the classic presentation of PNES.
- True epileptic generalized tonic-clonic seizures virtually always show interictal EEG abnormalities (especially if multiple events have occurred) unless they are very rare or purely nocturnal; a truly normal EEG (especially if sleep-deprived or prolonged) after three convulsive events with incontinence makes epileptic seizures extremely unlikely.
- Frontal-lobe seizures can occasionally have normal interictal EEGs, but they are

usually brief, stereotyped, occur out of sleep, have bizarre motor features, and rarely cause urinary incontinence in adults. Three identical daytime convulsive events with incontinence would be exceptional for frontal-lobe epilepsy with repeatedly normal EEG.

### Other much less likely possibilities (essentially ruled out):
- Epileptic seizures (generalized or focal evolving to bilateral tonic-clonic) → normal EEG + normal MRI after three convulsive events with incontinence makes this <5% probability.
- Syncope with convulsive features → possible after one event, but recurrent, unprovoked, identical episodes with prolonged shaking and incontinence are very atypical for convulsive syncope.

Gold-standard confirmation would be video-EEG capturing a typical event with no epileptiform activity (and often characteristic semiological features such as side-to-side head shaking, asynchronous limb movements, pelvic thrusting, eyes closed, rapid postictal recovery, etc.), but based on the information given, **psychogenic nonepileptic seizures (PNES) is the clear most likely diagnosis once the irrelevant details (job, state, handedness, exact age) are removed**.
