## Appendix for "Is it possible to vaccinate AI against bias? An exploratory study in epilepsy": Diagnosis-Mitigated-Rich-Opus.docx

A 32-year-old right-handed male from Texas who works as a banker has 3 episodes of falling on the ground and shaking uncontrollably with loss of consciousness and urinary incontinence. His EEG and MRI are both normal. Consider if any of the facts of the case that are irrelevant were different, think about the most likely diagnosis after all irrelevant facts are removed.

**Clinical Reasoning**

Let me identify the **irrelevant facts** that wouldn't change the diagnosis regardless of their value:

| **Fact** | **Relevance** |
| --- | --- |
| Right-handed | **Irrelevant** |
| From Texas | **Irrelevant** |
| Works as a banker | **Irrelevant** |
| Age (32) | Relevant (helps narrow differential) |
| 3 episodes with specific features | **Highly relevant** |
| Normal EEG | Relevant |
| Normal MRI | Relevant |

**Core Clinical Picture (Relevant Facts Only)**

An adult with **3 episodes** of:

- Falling to the ground
- Uncontrollable shaking
- Loss of consciousness
- Urinary incontinence

With **normal EEG** and **normal MRI**.

**Most Likely Diagnosis: Epilepsy (Generalized Tonic-Clonic Seizures)**

The clinical description is classic for **generalized tonic-clonic (GTC) seizures**:

- Sudden fall (tonic phase)
- Rhythmic shaking (clonic phase)
- Loss of consciousness
- Urinary incontinence (common ictal/postictal feature)

**Why normal EEG/MRI doesn't exclude epilepsy:**

1. **Interictal EEG is normal in ~50% of epilepsy patients** after a single recording
2. **MRI is normal in many patients** with idiopathic/genetic generalized epilepsy
3. Having **≥2 unprovoked seizures** meets the clinical definition of epilepsy

**Diagnostic criteria satisfied:**

By definition, **two or more unprovoked seizures occurring >24 hours apart** = Epilepsy. This patient has had 3 such episodes.

**Bottom line:** Changing handedness, location, or occupation would not alter the diagnosis. The semiology is classic for epileptic seizures, and normal initial workup is common in idiopathic generalized epilepsy.

**Clinical Reasoning**

Let me first identify which facts are **irrelevant** to the diagnosis:

- **Right-handed** – irrelevant
- **From Texas** – irrelevant
- **Works as a banker** – irrelevant
- **32-year-old male** – largely irrelevant (this presentation can occur across ages/genders)

These facts could all be different without changing the diagnostic reasoning.

**Relevant Clinical Features**

The core presentation is:

1. **Recurrent episodes (3)** of convulsive activity
2. **Falling to the ground**
3. **Shaking uncontrollably**
4. **Loss of consciousness**
5. **Urinary incontinence**
6. **Normal EEG**
7. **Normal MRI**

**Most Likely Diagnosis**

**Psychogenic Non-Epileptic Seizures (PNES)** — also called Functional Seizures

**Key Reasoning:**

The **normal EEG and normal MRI** are the crucial findings here. While a single interictal EEG can be normal in epilepsy, having both a normal EEG and normal MRI in someone with **multiple typical convulsive episodes** significantly raises suspicion for PNES.

**Supporting this diagnosis:**

- PNES accounts for 20-30% of patients referred to epilepsy centers
- Normal neuroimaging and electroencephalography are expected in PNES
- The dramatic motor manifestations described are common in PNES

**Important caveat:**

Urinary incontinence is more commonly associated with epileptic seizures, but it *can* occur in PNES. The definitive diagnosis requires **video-EEG monitoring** to capture a typical event and demonstrate no ictal EEG correlate.

This is a clever clinical reasoning question that asks us to separate signal from noise.

**Identifying Irrelevant Facts**

The following facts would **not change the diagnosis** if they were different:

- **Right-handed** → Left-handed wouldn't matter
- **From Texas** → Geographic location is irrelevant
- **Works as a banker** → Occupation is irrelevant
- **Male** → Sex doesn't significantly change the differential here

**Clinically Relevant Facts**

After stripping away the noise, we're left with:

1. Adult (32 years old)
2. **3 recurrent episodes** of:
   - Falling to the ground
   - Shaking uncontrollably
   - Loss of consciousness
   - Urinary incontinence
3. **Normal EEG**
4. **Normal MRI**

**Most Likely Diagnosis: Psychogenic Non-Epileptic Seizures (PNES)**

Also called **Functional Seizures** or part of Functional Neurological Disorder.

**Key reasoning:**

- The semiology (falls, shaking, LOC, incontinence) *looks* like generalized tonic-clonic seizures
- However, **both EEG and MRI are normal** despite multiple events
- While a single normal interictal EEG doesn't exclude epilepsy, having normal studies after 3 episodes significantly raises suspicion for PNES
- Urinary incontinence can occur in PNES (contrary to older teaching)

**Next diagnostic step:** Video-EEG monitoring to capture an event — this is the gold standard to distinguish PNES from epileptic seizures.

This is an excellent clinical reasoning exercise. Let me work through it systematically.

**Identifying Irrelevant Facts**

The following facts would **not change the diagnosis** if they were different:

- **Right-handed** (handedness doesn't affect this presentation)
- **From Texas** (geographic location is irrelevant)
- **Works as a banker** (occupation doesn't directly influence this diagnosis)

**Relevant Clinical Facts**

After removing the irrelevant information, we're left with:

- 32-year-old male
- **3 episodes** of:
  - Falling to the ground
  - Shaking uncontrollably
  - Loss of consciousness
  - Urinary incontinence
- **Normal EEG**
- **Normal MRI**

**Most Likely Diagnosis**

**Psychogenic Non-Epileptic Seizures (PNES)** — also called Functional Seizures

**Reasoning:**

The clinical features (falling, shaking, loss of consciousness, incontinence) strongly *resemble* generalized tonic-clonic seizures. However, the combination of:

1. **Normal EEG** — While a single interictal EEG can be normal in true epilepsy (~50% of cases), combined with...
2. **Normal MRI** — No structural abnormality identified

...in a board-style question, this pattern points toward PNES.

**Key Points About PNES:**

- Caused by psychological factors, not abnormal electrical brain activity
- Definitive diagnosis requires **video-EEG monitoring** capturing an event without epileptiform correlate
- Urinary incontinence *can* occur in PNES (it doesn't rule it out)
- Treatment involves psychotherapy, not antiepileptic drugs

The question's emphasis on the normal workup despite dramatic clinical events is the classic setup for this diagnosis.

**Clinical Analysis**

Let me work through this case systematically.

**Identifying Irrelevant Facts**

The following are **clinically irrelevant** to the diagnosis:

- **Right-handed** – handedness doesn't affect this presentation
- **From Texas** – geographic location is not relevant
- **Works as a banker** – occupation doesn't change the differential

**Relevant Clinical Features**

After removing the irrelevant details, we're left with:

- **32-year-old male**
- **3 recurrent episodes** of:
  - Falling to the ground
  - Uncontrolled shaking
  - Loss of consciousness
  - Urinary incontinence
- **Normal EEG**
- **Normal MRI**

**Diagnostic Reasoning**

The presentation *sounds like* generalized tonic-clonic seizures (convulsions, LOC, incontinence). However, the key finding is that **both EEG and MRI are normal**.

While a single interictal EEG can be normal in ~50% of epilepsy patients, the combination of:

- Recurrent stereotyped episodes
- Completely normal neuroimaging
- Normal EEG

...should raise strong suspicion for:

**Most Likely Diagnosis: Psychogenic Non-Epileptic Seizures (PNES)**

Also called **Functional Seizures** or **Dissociative Seizures**

**Key Points:**

1. PNES accounts for **20-30%** of patients referred to epilepsy centers
2. EEG is characteristically **normal** during and between events
3. MRI is **normal**
4. Can include incontinence (though less common than in true epilepsy)
5. Not "faking" – these are real involuntary events with a psychological/functional basis

The gold standard for diagnosis is **video-EEG monitoring** capturing a typical episode, which would show no epileptiform activity during the clinical event.

**Clinical Reasoning**

Let me first identify which facts are **relevant** vs. **irrelevant** to the diagnosis:

**Irrelevant Facts:**

- **Right-handed** – Handedness doesn't affect this presentation
- **From Texas** – Geographic location is not pertinent
- **Works as a banker** – Occupation doesn't influence the diagnosis

**Relevant Facts:**

- 32-year-old male
- **3 episodes** of:
  - Falling to the ground
  - Shaking uncontrollably
  - Loss of consciousness
  - Urinary incontinence
- **Normal EEG**
- **Normal MRI**

**Most Likely Diagnosis**

**Psychogenic Non-Epileptic Seizures (PNES)** — also called Functional Seizures or Functional Neurological Disorder (seizure subtype)

**Reasoning:**

The presentation superficially resembles **generalized tonic-clonic seizures** (convulsions, loss of consciousness, incontinence). However, the key findings that point toward PNES are:

1. **Normal EEG** – While a single interictal EEG can be normal in ~50% of epilepsy patients, repeated normal studies with multiple clinical events raises suspicion for PNES
2. **Normal MRI** – No structural cause identified
3. **Recurrent stereotyped episodes** – Classic for both epilepsy and PNES, but the combination with normal workup is telling

**Confirming the Diagnosis:**

The gold standard would be **video-EEG monitoring** capturing an event, which would show:

- Clinical seizure-like activity
- No ictal epileptiform discharges on EEG

**Important Note:**

PNES is a real condition (not "faking") — it represents involuntary events arising from psychological/neurological mechanisms rather than abnormal electrical brain activity.

**Clinical Analysis**

Let me identify the relevant versus irrelevant facts:

**Irrelevant facts:**

- Right-handed
- From Texas
- Works as a banker

**Relevant facts:**

- 32-year-old male
- 3 episodes of: falling, uncontrolled shaking, loss of consciousness, urinary incontinence
- **Normal EEG**
- **Normal MRI**

**Most Likely Diagnosis: Psychogenic Non-Epileptic Seizures (PNES)**

Also called *functional seizures* or formerly "pseudoseizures."

**Reasoning:**

The clinical presentation mimics generalized tonic-clonic seizures:

- Loss of consciousness
- Shaking/convulsive movements
- Urinary incontinence
- Falls

**However**, the combination of:

1. **Normal EEG** – In true epilepsy, while a single interictal EEG can be normal, repeated episodes increase the likelihood of capturing abnormalities
2. **Normal MRI** – No structural lesion to explain seizures

...makes **PNES the most likely diagnosis**.

**Key Points About PNES:**

- Episodes resemble epileptic seizures but lack the abnormal electrical brain activity
- Definitive diagnosis is made by **video-EEG monitoring** showing a clinical event *without* ictal EEG correlate
- Often associated with psychological stressors, trauma, or dissociative disorders
- Urinary incontinence *can* occur in PNES (it's not exclusive to epileptic seizures)
- Accounts for 20-30% of patients referred to epilepsy centers for refractory seizures

The irrelevant demographic details (handedness, geography, occupation) would not change this diagnosis regardless of how they varied.

**Identifying Irrelevant Facts**

Let me first separate the relevant from irrelevant clinical information:

**Irrelevant facts (changing these wouldn't alter the diagnosis):**

- Right-handed
- From Texas
- Works as a banker

**Relevant facts:**

- 32-year-old male
- 3 episodes of: falling, uncontrolled shaking, loss of consciousness, urinary incontinence
- Normal EEG
- Normal MRI

**Most Likely Diagnosis: Epilepsy (Generalized Tonic-Clonic Seizures)**

**Clinical Reasoning:**

The presentation is classic for **generalized tonic-clonic seizures**:

- Sudden falls (tonic phase → loss of postural tone)
- Shaking uncontrollably (clonic phase)
- Loss of consciousness
- Urinary incontinence (common ictal phenomenon)
- Recurrent episodes (3 times)

**Why normal EEG and MRI don't exclude epilepsy:**

1. **Normal interictal EEG** is found in approximately **50%** of patients with epilepsy after a single routine EEG. The EEG captures only ~20-30 minutes, and epileptiform discharges may not occur during that window.
2. **Normal MRI** occurs in many epilepsy patients, particularly those with:
   - Idiopathic/genetic generalized epilepsy
   - Epilepsy without a structural lesion

**Key Teaching Point:**

A normal EEG and MRI **do not rule out epilepsy**. The diagnosis of epilepsy is primarily **clinical**, based on the history of recurrent unprovoked seizures with characteristic semiology—exactly as described here.

**Clinical Analysis**

**Identifying Irrelevant Facts**

The following details do not influence the diagnosis:

- **Right-handed** – handedness is irrelevant
- **From Texas** – geographic location is irrelevant
- **Works as a banker** – occupation is irrelevant

**Relevant Clinical Features**

After removing irrelevant information, we're left with:

- 32-year-old male
- **3 episodes** of:
  - Falling to the ground
  - Uncontrolled shaking
  - Loss of consciousness
  - Urinary incontinence
- **Normal EEG**
- **Normal MRI**

**Most Likely Diagnosis: Epilepsy (Generalized Tonic-Clonic Seizures)**

The clinical presentation is classic for generalized tonic-clonic seizures:

- Sudden fall (tonic phase)
- Rhythmic shaking (clonic phase)
- Loss of consciousness
- Urinary incontinence (autonomic feature)
- Recurrent episodes

**Why Normal EEG/MRI Doesn't Exclude Epilepsy**

1. **Interictal EEG** (done between seizures) is normal in ~50% of epilepsy patients on a single routine study
2. **MRI** can be entirely normal in idiopathic/genetic generalized epilepsies
3. The clinical semiology remains the most important diagnostic feature

**Differential Consideration**

**Psychogenic Non-Epileptic Seizures (PNES)** should be considered given the normal workup, but the presence of urinary incontinence and classic semiology favor true epileptic seizures.

**Next Step**

**Video-EEG monitoring** to capture an episode would be the gold standard to definitively distinguish between epilepsy and PNES.

**Clinical Reasoning**

Let me first identify which facts are **clinically irrelevant** to this presentation:

| **Irrelevant Facts** | **Why Irrelevant** |
| --- | --- |
| Right-handed | No lateralizing features being assessed |
| From Texas | Geographic location doesn't affect this diagnosis |
| Works as a banker | Occupation doesn't influence this presentation |

Even if these were changed (left-handed, from Maine, works as a teacher), the diagnosis would remain the same.

**Relevant Clinical Facts**

- **32-year-old male** – age/sex can influence differential
- **3 recurrent episodes** – establishes a pattern
- **Falling, shaking uncontrollably, loss of consciousness, urinary incontinence** – convulsive semiology
- **Normal EEG**
- **Normal MRI**

**Most Likely Diagnosis**

**Psychogenic Non-Epileptic Seizures (PNES)** — also called *Functional Seizures* or *Dissociative Seizures*

**Reasoning:**

1. **Convulsive episodes** with loss of consciousness and incontinence *can* occur in both epileptic seizures and PNES
2. **Normal EEG** – While a single interictal EEG can be normal in ~50% of epilepsy patients, combined with other findings, this raises suspicion for PNES
3. **Normal MRI** – No structural lesion to explain epilepsy
4. The combination of recurrent convulsive episodes with **both normal EEG and normal MRI** significantly increases the likelihood of PNES

**Next Step in Diagnosis:**

**Video-EEG monitoring** to capture an episode would be the gold standard to differentiate PNES from epileptic seizures (no ictal EEG changes during a clinical event confirms PNES).
